# Network-Based Methods for Psychometric Data of Eating Disorders: A Systematic Review

**DOI:** 10.1101/2022.03.15.22272402

**Authors:** Clara Punzi, Manuela Petti, Paolo Tieri

## Abstract

Network science represents a powerful and increasingly promising method for studying complex real-world problems. In the last decade, it has been applied to psychometric data in the attempt to explain psychopathologies as complex systems of causally interconnected symptoms. With this work, we aimed to review a large sample of network-based studies that exploit psychometric data related to eating disorders (ED) trying to highlight important aspects such as core symptoms, influences of external factors, comorbidities, and changes in network structure and connectivity across both time and subpopulations. A particular focus is here given to the potentialities and limitations of the available methodologies used in the field. At the same time, we also give a review of the statistical software packages currently used to carry out each phase of the network estimation and analysis workflow. Although many theoretical results, especially those concerning the ED core symptoms, have already been confirmed by multiple studies, their supporting function in clinical treatment still needs to be thoroughly assessed.

## Introduction

In the last century, the paradigm that best got ahead in Western medicine has been the “disease model” (Borsboom & Cramer, 2013), according to which all symptoms a person exhibits result from a latent entity that should therefore be targeted to an effective treatment to obtain, as a consequence, the lessening of all the deriving symptoms (Jones et al., 2017; McNally, 2021).

Unfortunately, in contrast with general medicine, in most mental disorders the identification of common pathogenic pathways has proven elusive (Borsboom, 2017; Borsboom & Cramer, 2013; McNally, 2016, 2021), given that they cannot be diagnosed independently of their symptoms (Borsboom & Cramer, 2013). Therefore, the need of conceptualizing in an alternative way the relation between symptoms and disorders arose in the twenty-first century and led to the delineation of the network theory of psychopathology, an innovative approach that inspired an exponentially increasing number of empirical publications in the past two decades, especially after the seminal article by Borsboom and Cramer (2013) was published. Differently from the disease model, in the network model, symptoms are conceptualized as mutually interacting and reciprocally reinforcing elements of a complex network, i.e., causally active components of the mental disorder instead of passive receptors of its causal influence (see Figure 1).

**Figure 1.**
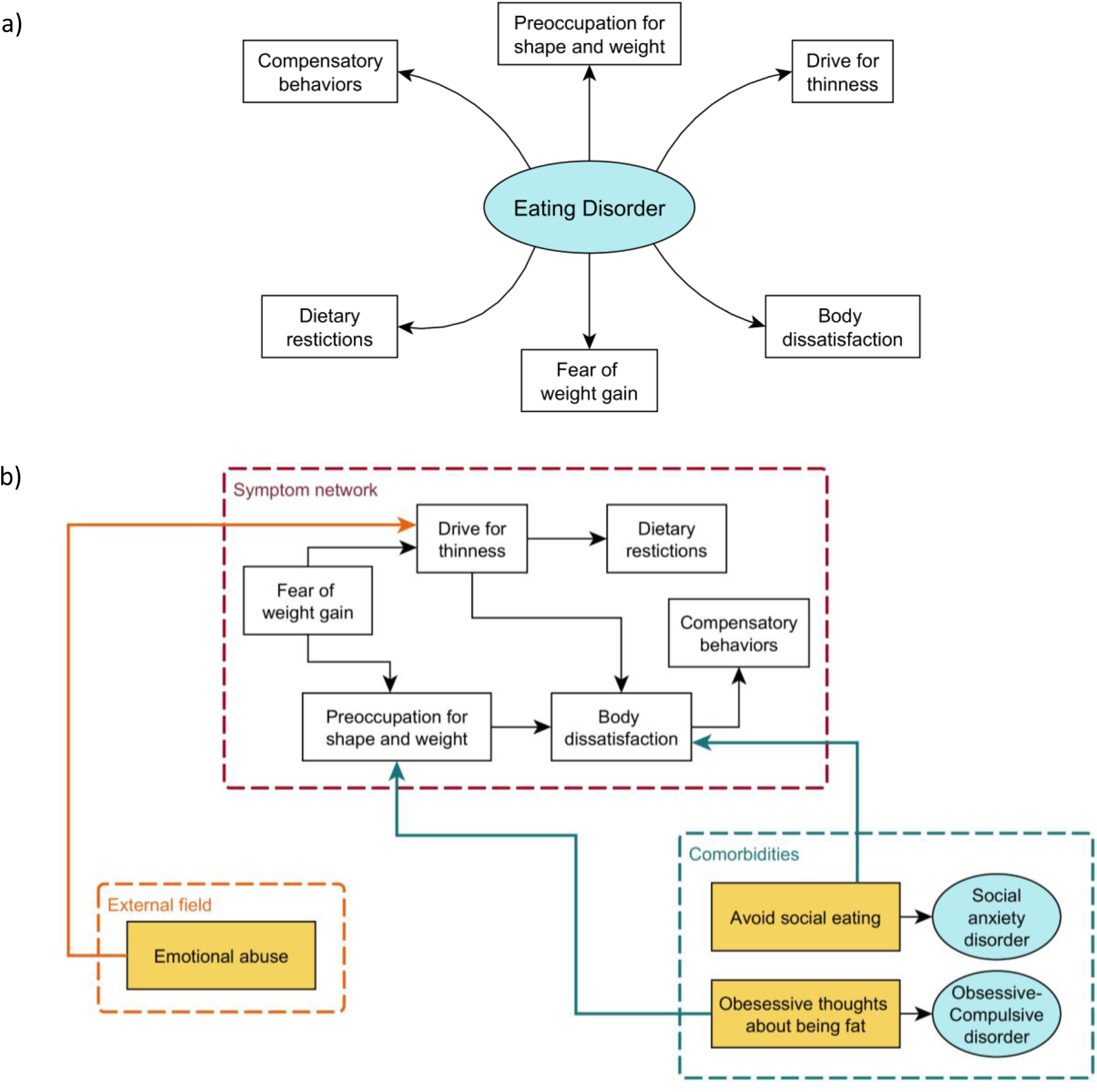
Comparison between Factor and Network Model. Schematic representation of factor model (a) and network model (b) of eating disorders (simplified). While in the first case symptoms (white rectangles) are considered manifestations of some common underlying factor (e.g., the eating disorder psychopathology [cyan ellipse]), according to the network model symptoms are conceptualized as mutually interacting and reciprocally reinforcing elements of a complex network where ED-specific symptoms (white rectangles in the red dashed box) mutually influence non-specific ones (yellow rectangles), such as external events (orange dashed box) or comorbidities (cyan ellipses in the blue dashed box). Hence, symptoms are seen as causally active components of the mental disorder instead of passive receptors of its causal influence.

In the present work, we aim to describe the current state-of-the-art of the network conceptualization of a specific psychopathology, namely eating disorders (EDs). These are severe psychiatric syndromes defined by abnormal eating behaviors that negatively affect a person’s physical or mental health (American Psychiatric Association, 2013). They are believed to result from and be sustained by sociocultural, psychological, and biological factors. Anorexia nervosa (AN), bulimia nervosa (BN), and binge eating disorder (BED) are the primary diagnoses associated with ED.

### The Network Approach to Psychopathology: a Theoretical Framework

The central idea behind the network approach to psychopathology is that mental disorders arise from casual interactions between symptoms, where causality must be interpreted in the sense of the interventionist theory, according to which the relation between two symptoms is causal if there exists a possible (natural or experimental) intervention on one of them that changes the probability distribution of the other, independently of the how the causal relationships are triggered (Borsboom, 2017).

In order to represent and study these symptom-symptom interactions, a network structure can be used, a so-called *symptom network*. In the scientific setting, the term *network* refers to a mathematical structure called *graph*, which consists of a set of nodes connected by links, or, in more formal terms, an ordered pair *G=(V, E)* where *V* is the set of vertices (or nodes), i.e., the system’s components, and *E* is the set of edges (or links), i.e., the interactions between them. If the edges have no direction, thus indicating a two-way relationship, then the graph is said to be *undirected*; otherwise, if the edges are given a specific direction, that is, they can only be traversed in a single direction, then the graph is called *directed*. Moreover, each edge can also be given a number called *weight*, that represents a quantification of its strength or cost or capacity, according to the different context. In this case, the graph is said to be *weighted* to distinguish it from the *unweighted* type (Bollobás, 1998). The arrangement of the network’s elements is called *topology*.

Although no distinction is usually made, the terminology “graph”, “vertex”, “edge” refers more precisely to the mathematical representation of the system, whereas “network”, “node”, “link” is more common in reference to real systems such as physical, biological, social, and economical systems. The network approach has proven successful and useful in a number of fields, from social sciences to economics, informatics, ecology, epidemics, biology, and medicine, among others (cfr. Barabási 2004; 2011; 2016; Tieri et al., 2019). In the specific case of symptom networks, nodes encode symptoms and edges stand for causal influence between pairs of symptoms.

There might also be conditions that can change the state of symptoms from the outside of the psychopathology network, for example adverse life events, abnormal brain functioning and inflammation, among others; all together, these constitute the *external field* of the symptom network (Borsboom, 2017). Another crucial property is the existence of partially overlapping syndromic clusters or bridge symptoms, that is, symptoms that are associated with multiple disorders and thus are part of symptom networks corresponding to different psychopathologies. This feature allows for an immediate explanation of the high level of comorbidity that characterizes mental disorders (Borsboom, 2017; Cramer et al., 2010; McNally, 2016, 2021).

An ultimate facet that needs to be considered to complete the theoretical framework of network analysis applied to psychopathology is the proven existence (in most psychopathology networks) of a phenomenon called *hysteresis*, which is a fundamental indicator of the dynamics of the system and consists in the dependence of any state of the system to its history (Cramer, 2013). In other words, once a system has been activated by an external event, the subsequent fading of that event will not necessarily deactivate that symptom in case there exist connections with other symptoms that are strong enough to make the reactions provoked by the triggering event (i.e., the activated symptoms system) self-sustaining (Borsboom, 2017). As proven by Cramer (2013) the hysteresis effect becomes more pronounced as the connectivity of a network increases. In fact, what has been noticed is that in weakly connected networks, even though significant triggering events can cause strong reactions, once the event is over, the system will gradually recover and return to its asymptomatic state. In this sense, a weakly connected network is said to be *resilient*, as opposed to the *vulnerable* disposition of the strongly connected ones (Borsboom, 2017).

Following these observations, Borsboom (2017) proposed new definitions of *mental health* as the stable state of a weakly connected network and *mental disorder* as an alternative stable state of a strongly connected network which is separated from the healthy state by hysteresis.

At this point, it is important to underlie that the conceptualization of the network approach to mental disorders should not be regarded as a theoretical finding only. Indeed, it has remarkable implications for the diagnosis and treatment systems as well (Borsboom, 2017).

### The Psychometric Network Analysis Workflow

The term *psychometric network analysis* is used to describe the combined procedure of network estimation, network description and network stability analysis, which together build the bulk of the methodology used in network approaches to multivariate psychometric data (Borsboom & Cramer, 2013; Borsboom et al., 2021; Christensen et al., 2018; Cramer et al., 2010; Forbes et al., 2017).

The complete workflow (Figure 2) typically starts with a specific research question, according to which a suitable data collection scheme is chosen. Usually, experimental data is given in the form of either a cross-sectional, time-series or panel design. Although the subsequent procedures are generic statistical ones and thus apply to input variables of any kind, in this context psychometric variables usually consist of responses to questionnaire items, symptom ratings and cognitive test scores together with other possible personal or psychological indicators (Borsboom et al., 2021).

**Figure 2.**
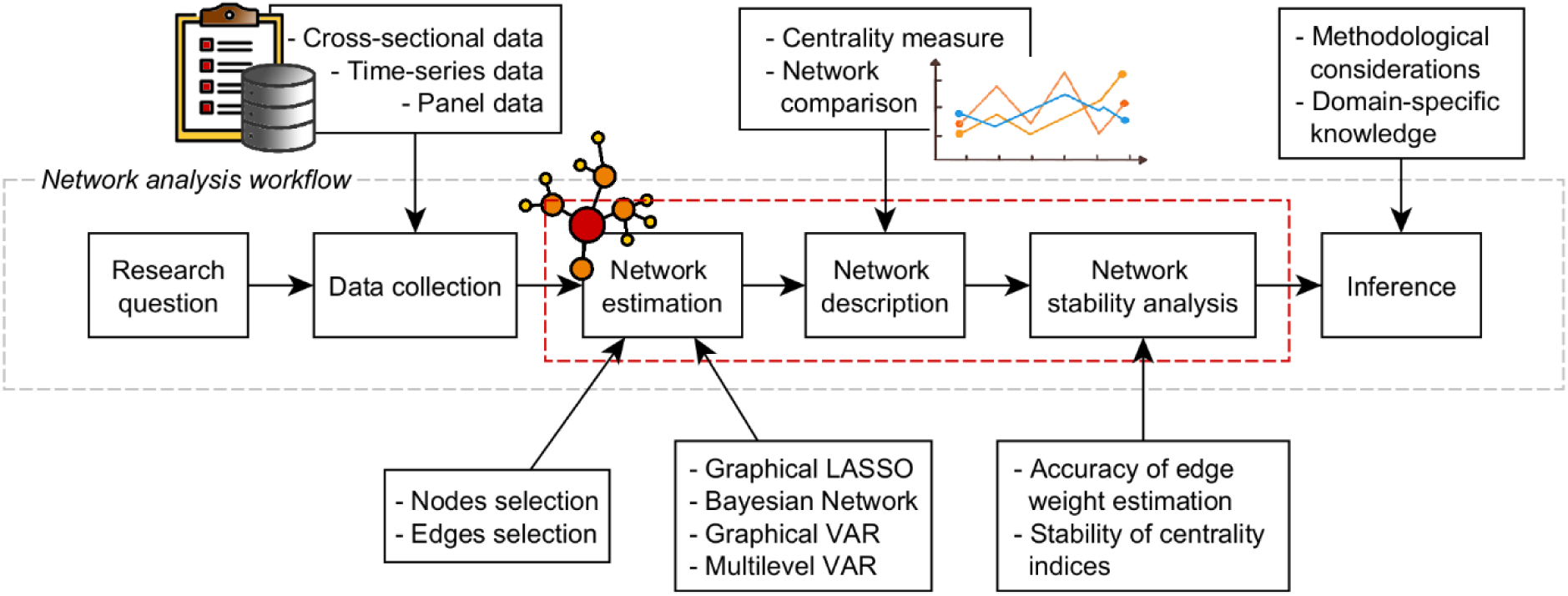
**Psychometric Network Analysis Workflow**. Scheme of the typical workflow of psychometric network analysis. Once the research question has been defined (also according to the availability of data), the main steps to be performed are: 1. Network estimation, that is, construction of the network. 2. Network description, that is, identification of important symptoms. 3. Network stability analysis, that is, assessment of the robustness of results. Together, these will allow to infer significant interpretation that should be employed in clinical treatment.

Once enough data are available, the network estimation step can be carried out with the aim of approximating the values of links between pairs of nodes (i.e., the causal influence of one onto the other) and building an appropriate network structure at the system level. Depending on the peculiarities of the data, different statistical methods can be employed: the most frequent approach is that of assessing the edge parameters as conditional associations between variables to estimate the corresponding *Pairwise Markov Random Field* (PMRF), but the alternative strategy of *Bayesian network* estimation has been successfully employed as well (McNally, 2021). Importantly, this step also encompasses the process of *node selection* and *edge selection*, the latter via general statistical methods such as fit indices, null-hypothesis testing, or cross-validation procedures.

The result of this step is generally a nontrivial topological structure which becomes the main subject of the *network description* phase, whose aim is to give a complete characterization of the symptom network with a particular focus on its most important nodes. Here, “importance” has to be intended as how a node is interconnected with the other nodes of the network and is commonly assessed by different centrality measures (McNally, 2021), that is, scalar values assigned to each node within a graph in order to assess their significance based on certain definitions of importance. In general, the tools of network analysis are employed to estimate network density and connectivity through global topological properties, node centrality through local topological properties and more fine-grained structural patterns such as communities and motifs (i.e., mesoscopic level; Letina et al., 2018).

Next, it is fundamental to evaluate the stability and robustness of the estimated network and of the centrality indices. In fact, the estimation error and the sampling variation need to be considered in order to not obtain misleading results (Borsboom et al., 2018; Christensen & Golino, 2021; Epskamp, Borsboom, et al., 2018; Fried & Cramer, 2017; Fried et al., 2021). Altogether, the methods used to assess the accuracy of the estimated parameters and their ability to replicate in a different dataset constitute the *network stability analysis* (Fried & Cramer, 2017).

Finally, the psychometric network analysis approach comes to an end with proper inferences which require taking into account both substantive domain knowledge and methodological considerations about the stability and robustness of the estimated network (Borsboom et al., 2021).

The aim of this study is to review the existing literature on psychometric network analysis of EDs. To our knowledge, three other reviews on EDs have already been published (Levinson et al., 2018c; Monteleone & Cascino, 2021; Smith et al., 2018). We intend to update and broaden the results of such seminal reviews with a larger and wide-ranging sample of studies, in particular by focusing on the potentialities and limitations of the available methodologies in the field of psychometric network analysis.

## Methods

To ensure a standardized review procedure, the Preferred Reporting Items for Systematic reviews and Meta-analyses (PRISMA) 2020 statement (Page et al., 2021) was followed. The articles included in this study were extracted from those returned by the query run on PubMed in February 2022 with search keys *((“eating disorder*”[Title/Abstract]) OR (“bulimia nervosa”[Title/Abstract]) OR (“anorexia nervosa”[Title/Abstract])) AND (network analys*[Title/Abstract])*. The original collection of 89 papers was further narrowed down to a number of 54 by considering the following exclusion criteria: (1) studies not based on psychometric data, (2) studies with aims different from the investigation of eating disorders, and (3) review articles. The final subset of 56 papers was obtained by adding two more articles cited as references in the selected publications. The list of such papers is provided in the supplemental materials. The PRISMA flow chart corresponding to the present methodology is shown in Figure 3.

**Figure 3.**
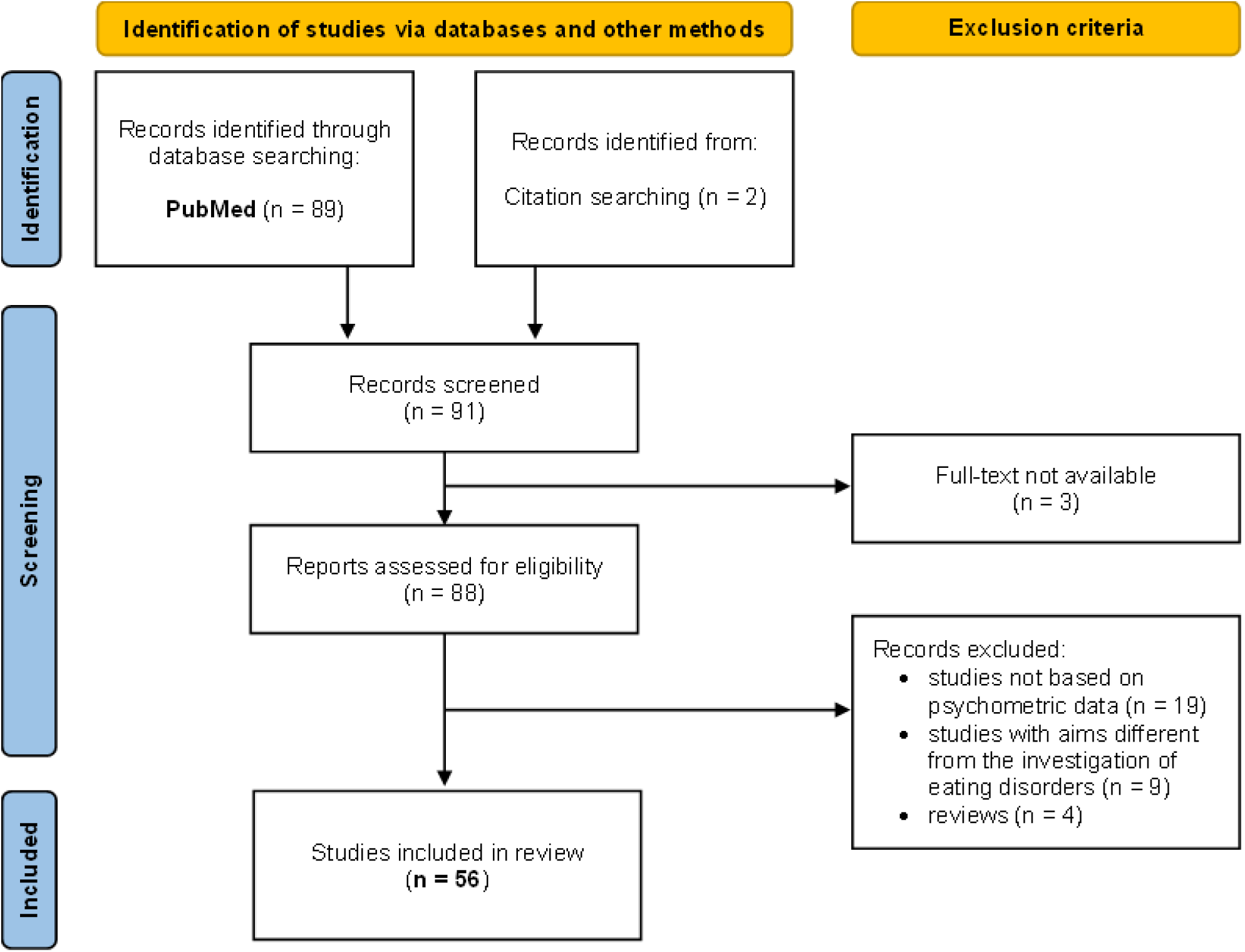
PRISMA flow chart for selection and filtering of studies.

## Results

In the following, we analyzed a large sample of network-based studies that exploit psychometric data related to ED. Specifically, we first introduce each step of the (general) psychometric network analysis workflow and then describe and compare the results reported in the articles under review.

### Research Question

In line with the application to other mental disorders, various research goals can be identified among the existing literature about network approaches to EDs, namely:

A. validation of the transdiagnostic model of eating disorders by comparing network characteristics across ED diagnoses (DuBois et al., 2017; Forrest et al., 2018; Goldschmidt et al., 2018; Mares et al., 2021; Monteleone, Tzischinsky, et al., 2022; Solmi et al., 2018; Solmi et al., 2019);
B. estimation of the symptom network of EDs and identification of the core symptoms (Beauchamp et al., 2021; Forbush et al., 2016; Forrest et al., 2018; Forrest, Perkins, et al., 2019; Rodgers et al., 2018; Wang et al., 2019);
C. identification and interaction with nonspecific ED symptoms (i.e., the external field) like general psychiatric symptoms, personality traits and other clinical variables (Monteleone, Mereu, et al., 2019; Solmi et al., 2018; Solmi et al., 2019), embodiment dimensions (Cascino et al., 2019), childhood maltreatment (Liebman et al., 2021; Monteleone, Cascino, et al., 2019; Monteleone, Tzischinsky, et al., 2022; Rodgers et al., 2019), mentalizing and empathy (Monteleone et al., 2020), vulnerability factors (Vervaet et al., 2021), suicidal thoughts and behaviors (Smith et al., 2020), perfectionism and interoceptive sensibility (Martini et al., 2021), affective and metacognitive symptoms (Aloi et al., 2021; Wong et al., 2021), interoceptive awareness (Brown et al., 2020), sleep disturbance (Ralph-Nearman et al., 2021), well-being domains (de Vos et al., 2021), inflexible and biased social interpretations, socioemotional functioning (Bronstein et al., 2022);
D. assessment of psychiatric comorbidities such as depression and anxiety (Bronstein et al., 2022; Elliott et al., 2020; Kenny et al., 2021; Levinson et al., 2017; Sahlan, Williams, et al., 2021; Smith et al., 2019), posttraumatic stress disorder (Liebman et al., 2021; Vanzhula et al., 2019), social anxiety disorder (Levinson et al., 2018a; Sahlan, Keshishian, et al., 2021), obsessive-compulsive disorder (Giles et al., 2022; Kinkel-Ram et al., 2021; Meier et al., 2020; Vanzhula et al., 2021), trait anxiety disorder (Forrest, Sarfan, et al., 2019), autism spectrum disorder (Kerr-Gaffney et al., 2020), borderline personality disorder (De Paoli et al., 2020), and alcohol misuse (Cusack et al., 2021);
E. comparison of estimated network structures among clinical and nonclinical (Vanzhula et al., 2019), ethnic minority women (Perez et al., 2021), men and women (Perko et al., 2019), across developmental stages (Calugi et al., 2020; Christian et al., 2020; Schlegl et al., 2021), and across different duration of illness (Christian et al., 2021);
F. characterization of the dynamic structure of systems and evaluation of intraindividual networks (Levinson et al., 2021; Levinson et al., 2018b; Levinson et al., 2020);
G. assessment of treatment outcome (Calugi et al., 2021; Elliott et al., 2020; Hagan et al., 2021; Hilbert et al., 2020; Olatunji et al., 2018; Smith et al., 2019).

### Collection of Psychometric Data

The accomplishment of the above research goals relies on the successful collection of datasets having specific peculiarities, since this will determine the possibility of estimating certain types of networks. The most typical starting point for this kind of analysis is clearly the selection of appropriate psychometric assessment tools, mainly self-report questionnaires and structured clinical interviews (McNally, 2021). Depending on the sample size and the sampling frequency, three types of data environments can be identified among the current practice of network approaches to psychopathology, namely cross-sectional data, time-series data, and panel or longitudinal data.

*Cross-sectional data* has been the first type of data used in this field and is definitely the most mentioned across the existing literature (e.g., it was used in 48 papers out 56 in our sample). It is particularly suitable for the estimation of group-level networks, since it provides variable measures taken at a single time point in a large sample. Importantly, the associations between variables are built upon differences among individuals and for such a reason a lot of caution should be taken when inferences about single patients are made, since the very strict conditions under which a structure of intraindividual variation can be deduced from the analogous structure of interindividual variation are seldom met in psychological processes (Bos & Wanders, 2016; Molenaar, 2009). In a cross-sectional dataset, rows can be reasonably assumed to be independent, therefore the corresponding PMRF can be directly estimated from it.

*Time-series* and *panel data* have been introduced in the psychometric network modeling in order to address two main limitations of cross-sectional data: the lack of clear understanding of individual networks and the inability to capture the dynamic features of psychopathology (McNally, 2021). Both data environments are characterized by datasets where variables are measured at multiple time points, with the difference that time-series data focuses on one single individual, whereas panel data consist of observations of multiple individuals. Given a time-series dataset, one can estimate two different structures: a directed temporal network of vector autoregressive coefficients where links describe associations between variables through time, and an undirected contemporaneous network where links describe instead the association between variables after the temporal effects have been removed. In case of panel data, a third structure can be estimated, namely a between-subject network, where links indicate the conditional associations between the long-term averages of the time series between people (Borsboom et al., 2021).

In line with other experimental studies, the applications to EDs mostly move from cross-sectional data. Nevertheless, few exceptions are worth mentioning. Firstly, Levinson and coworkers in three different papers (2021; 2018; 2020) used panel data to estimate interindividual networks (temporal, contemporaneous, and between-subject), as well as intra-individual networks (temporal and contemporaneous) for some of the patients in the sample. Other relevant studies aimed at assessing the treatment efficacy by applying statistical techniques (Brown et al., 2020; Calugi et al., 2021; Christian et al., 2020; Elliott et al., 2020; Hagan et al., 2021; Hilbert et al., 2020; Smith et al., 2019).

Data vary in terms of other features as well. Among the articles under review, 49 out of 56 described their sample as being composed of a great majority of female participants. After all, the fact that EDs are much more common in women than in men is broadly known and documented (Striegel-Moore et al., 2009), with reasons usually attributed to social pressure (Whiteman, 2016), adolescent turbulence, poor body concept, and role confusion (Hsu, 1989). Just one study involved only male participants (Forrest, Perkins, et al., 2019), while few other papers reported a more heterogeneous (mostly nonclinical) sample with male participants within the range of 40-60% (Bronstein et al., 2022; Kenny et al., 2021; Kinkel-Ram et al., 2021; Liebman et al., 2021; Perko et al., 2019; Sahlan, Keshishian, et al., 2021; Sahlan, Williams, et al., 2021).

Moreover, in 60% of the cases, participants are reported as clinical, either inpatients or outpatients. Among these, three studies involved users of the Recovery Record^1^ smartphone application (Christian et al., 2020; 2021; Perko et al., 2019). Exceptions consist in mixed samples involving nonclinical patients, such as school or college students, and three case studies based on datasets collected through the crowdsourcing marketplace Amazon Mechanical Turk, (MTurk; Forrest, Perkins, et al., 2019; Kinkel-Ram et al., 2021; Liebman et al., 2021).

Nearly all papers focus on the most common ED diagnosis, namely Anorexia Nervosa (AN) and Bulimia Nervosa (BN). However, some of them also present results concerning secondary EDs, in particular binge-eating disorder (Hilbert et al., 2020; Wang et al., 2019), and night eating disorders (Beauchamp et al., 2021),

Various psychometric assessment questionnaires were used as tools for data collection. For the evaluation of ED specific symptoms, the most widely used tests were the Eating Disorder Inventory (EDI; Garner et al., 1983; Garner, 1991; 2004), the Eating Disorder Examination Questionnaire (EDE-Q; Fairburn & Beglin, 1994), and the Eating Pathology Symptom Inventory (EPSI; Forbush et al. 2013). For the assessment of general psychological factors other tests were also used, for example the Symptom Check-List 90 (SCL-90; (Derogatis & Cleary, 1977; Derogatis & Unger, 2010), and the Beck Depression Inventory (BDI-II; Beck et al., 1996). Note that, among the cited psychometric tests, the only one that has been designed to assess both ED specific symptoms as well as other general integrative psychological constructs is EDI.

### Methods for Network Estimation and Reconstruction

Once the data has been collected, the next fundamental point is determining the variables of interest, that is, the nodes of the network. Instead of considering the totality of the items included in the questionnaires, a common practice is that of reducing their number in an effort to produce more accurate results by avoiding redundant (i.e., collinear) variables. The final nodes do not generally comprise all the items of the questionnaires. Instead, they are chosen in either one of the following ways, namely by taking into account just some special item aggregates such as questionnaires’ subscales, by employing the *goldbricker()* function of the *R* package *networktools* (Jones, 2018), which compares correlations between variables and identifies the collinear ones, or by combining the latter with a further theoretically driven selection of items.

#### Cross-Sectional Networks

The types of networks that can be estimated depend on what kind of data is available. In case of cross-sectional data, the main solutions are association networks, concentration (or partial correlation) graphs, regularized partial correlation networks, and Bayesian networks, where the first three types are undirected, weighted and can all be estimated with the *qgraph* (Epskamp et al., 2012) *R* package, while the last one is direct, either weighted or unweighted, and can be obtained with the help of the *bnlearn* (Scutari, 2009) *R* package.

*Association networks* are the most basic types of networks that can be estimated from cross-sectional data. Edges correspond to zero-order correlations between symptoms, indicating the probability of their co-occurrence. For example, the *qgraph() R* function with input parameter graph = “cor” will compute an association network by estimating zero-order correlations between each pair of variables through the Pearson coefficient *r*.

Consider a set of *p* variables 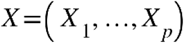, each described by *n* observation, that is,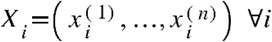. Given paired data 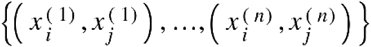, the Pearson coefficient

is defined as:

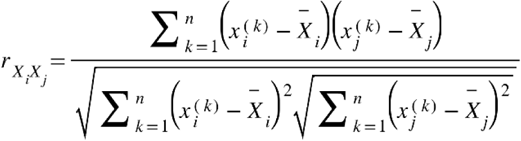

where *n* is the sample size, 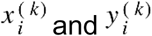 are two sample points indexed with *k*, and 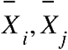 are the sample means of the variables 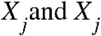.

Association networks have two main limitations: first, they do not give any information about the direction of causal relationships, and second, they do not discern true relations from spurious ones and from those caused by the influence of other nodes (McNally, 2021).

*Concentration networks* solve the second of these limitations by estimating edges as partial correlations between symptoms after adjusting for the influence of all other nodes in the networks; only the edges whose value is above a fixed threshold are then kept. Formally, the partial correlation between two variables *X* and *Y* given a set of *n* controlling variables 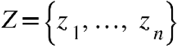 is written as 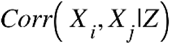 and is given by the correlation between the residuals *e_X_* and *e_Y_* resulting from the linear regression of *X* with *Z* and of *Y* with *Z*, respectively. A network where each edge corresponds to the partial correlation between the connected nodes can be estimated through the *qgraph()* function by setting the parameter graph = “concentration”.

When dealing with *p*-multivariate data 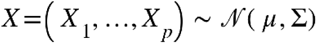 all information needed to compute the partial correlation coefficients is encoded in the variance-covariance matrix Σ. In fact, once its inverse is defined (i.e., the so-called precision matrix *K*), one can directly apply the following relationship to recover the partial correlation coefficients:

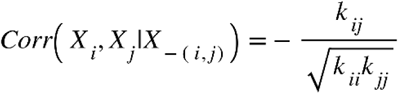

where *k_ij_* denotes the element in row *i* and column *j* of the precision matrix *K* = ∑^−1^ and 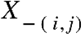 denotes the set of variables without *X_i_* and *X_j_*. These coefficients can be graphically displayed in a weighted undirected network where each node corresponds to a variable and edges between them are given by the partial correlation coefficients. If the *ij*^th^ component of ∑^−1^ is zero, then the variables *X_i_* and *X_j_*are conditionally independent, given the other variables, and no edge will be traced between them. This model is a type of PMRF and it is called *Gaussian Graphical Model*, shortly GGM (Epskamp, Borsboom, et al., 2018). Forbush et al. (2016) gave an example of an ED symptom network estimated as an association graph and also proposed the corresponding concentration network in the supplementary material of the same paper.

When the number of variables to estimate is high, it has been suggested that a more appropriate model to use is the *regularized partial correlation network* (Epskamp & Fried, 2018), obtainable running the *gqraph() R* function with input parameter graph = ”glasso”. The result is similar to a concentration graph in the fact that edges indicate partial correlation between nodes, however it has the relevant difference of including the implementation of an L1 regularization technique called *graphical LASSO* (least absolute shrinkage and selection operator; Tibshirani, 1996) which allows for the shrinking of all small partial correlations to zero. This procedure returns a sparse network that parsimoniously accounts for the covariance among nodes, in the sense that only the edges that are most robust and most likely to reflect real associations are kept (McNally, 2021). Formally, the graphical LASSO gives an estimation of the precision matrix *K* = ∑^−1^ by solving the optimization problem of maximize the penalized log-likelihood

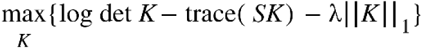

over nonnegative definite matrices *K*, where *S* is the empirical covariance matrix of *X*, λ is a nonnegative tuning parameter and ||*K*||_1_ denotes the *ℓ*-norm, that is, the sum of the absolute values of the elements of ∑^−1^. Hence, the higher the λ value, the more *K_ij_* will be set to zero.

Clearly, if λ is too low, then too many spurious edges risk being included (i.e., yielding a high number of false positives), whereas if λ is too high, then the risk is to remove relevant connections (false negatives). Hence, the λ parameter needs to be tuned. The best model (i.e., the most likely to maximize the number of “true” edges while minimizing the spurious ones) is then identified through the Extended Bayesian Information Criterion (EBIC; Chen & Chen, 2008) for multiple λ values and then choosing the model with lowest EBIC score. Let *P* be a subset of {1, …*p*} and let *ν* = |*P*| be the cardinality of this subset, then the best λ is chosen to be the one that maximizes

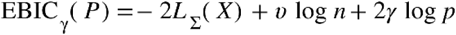

where 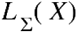 is the log-likelihood 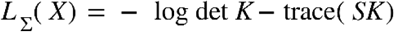 (Chen & Chen, 2008; van Bork et al., 2018).

It has been suggested by Williams and Rast (2020) that the graphical LASSO gives an accurate representation of data only when the number of nodes vastly exceeds the number of cases; if not, a nonregularized network should be preferred. Nevertheless, all except one of the articles under review based on cross-sectional data employ the graphical LASSO to estimate the symptom network.

*Bayesian networks* attempt to discern causality by representing data as directed acyclic graphs (DAGs) where arrows indicate the direction of predictions and, possibly, causality (McNally, 2016). DAGs depict the joint probability distribution of the variables and can thus be decomposed into the conditional distribution of each node given its parent. Importantly, dependence relations should not be confused with temporal antecedence, which cannot be derived from cross-sectional data in any way. What restrains Bayesian networks from widespread application is the existence of strict assumptions about data that it is pretty difficult to find in psychological analysis settings according to clinical observations (McNally, 2021). The first condition is that all relevant causal variables should be included in the system. Second, the causal Markov Condition^2^ (McNally 2016) must be met. Third, the probability distributions of certain variables might not be unrestricted. And fourth, it might not be easy to choose the best model among all possible ones (McNally, 2016). Moreover, one additional assumption is suggested by the definition itself of DAGs, namely that all loops of any length are prohibited. A Bayesian network can be estimated from multivariate data through the *bnlearn* R package (Scutari, 2009).

In a paper that analyzes the relationship between EDs and childhood abuse, Rodgers et al. (2019) followed this Bayesian approach in two steps: they first wrote down a “blacklist” of forbidden edges to limit the investigation to patterns of symptom relationships that made both conceptual and clinical sense; next, they estimated the DAG through the hill-climbing algorithm. This is an iterative machine-learning process that starts with an arbitrary network structure and tries to improve it by making incremental changes to the network (e.g., adding, removing, or reversing edges). At each step, the Bayesian Information Criterion (BIC) is computed, and the change is kept if and only if it results in a lower BIC. This process continues until no further improvements can be found and the network model that represents the best fit for the interactive structure of the ED psychopathology is returned.

Diverging from the other studies, Bronstein et al. (2022) employed a combined procedure to test different hypotheses. In particular, they used the exploratory causal discovery algorithm *Greedy Fast Causal Inference* (GFCI; Ogarrio et al., 2016) to investigate potential causal pathways involving ED symptoms, biased and inflexible interpretations, and socioemotional functioning. GFCI takes as input a dataset of continuous variables (e.g., psychometric data) and outputs a graphical model called *Partial Ancestral Graph* (PAG; Richardson, 1996), which is a representation of a set of Bayesian Networks that cannot be distinguished by the algorithm. Notably, GFCI is characterized by the ability of detecting latent confounders^3^; this information is conveyed by different edge types in the generated PAG.

#### Joint Estimation of Cross-Sectional Networks

In many of the studies under review, multiple networks were estimated with the specific aim of comparing their structure in various populations. As it will be later explained in more details, most often this task is accomplished by first estimating each network separately and only later some pivotal test statistics are computed to highlight global and local topological differences.

However, when the observations in a dataset consist of several distinct classes, it is also possible to adopt a recently developed technique called *Joint Graphical Lasso* (JGL), which allows for jointly estimating multiple graphical models corresponding to distinct but related conditions (Danaher et al., 2014). In particular, the JGL fits GGMs on data with the same variables observed on different classes but differs in the estimation of uncoupled GGMs from independent samples in the fact that, besides the lasso penalty on density, the regularized optimization problem for the evaluation of the precision matrices of each class includes an additional penalty term that is specifically written to foster similarity between groups.

Suppose we are given *K* datasets 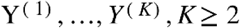, where each *Y*^(*k*)^ is a 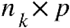 matrix consisting of *^n^_k_* observations on a set of *p* features common to all *K* datasets. We assume that all the observations 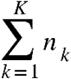 are independent and that, within each dataset, 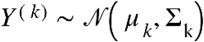. Then one can define the empirical covariance matrix for*Y*^(*k*)^as 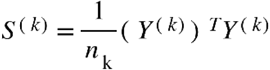. Danaher et al. (2014) proposed to estimate the precision matrices 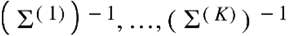 by maximizing the penalized log-likelihood

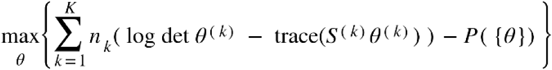

where 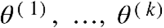 are assumed to be positive definite and *P*({*θ*}) denotes a convex penalty function chosen to encourage precision matrices to share certain characteristics (e.g., the locations or values of the nonzero elements or the sparsity).

Depending on the explicit definition of the penalty function, JGLs are classified as Fused Graphical Lasso (FGL) and Group Graphical Lasso (GGL). The former encourages shared edge values across classes, whereas the latter only encourages a similar pattern of sparsity across all precision matrices. Hence, the FGL results in a stronger form of similarity (Danaher et al., 2014). The penalty of the FGL has the form

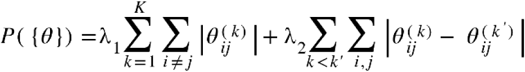

where *λ*_1_ and *λ*_2_ are nonnegative tuning parameters, the first controlling thè*ℓ*-penalties applied to each off-diagonal element of the *K* precision matrices, the second those applied to the differences between corresponding elements of each pair of precision matrices. Hence, large values of *λ*_1_ will increase the sparsity of the precision matrices (just like in graphical LASSO), while large values of *λ*_2_ will cause many elements to be identical across classes.

Both types of JGL have already been implemented in the *R* package *EstimateGroupNetwork* (Costantini et al., 2019), which also include methods for the automatic tuning parameter selection. A final consideration about JGL is that its network estimations cannot behave worse than independent GGMs (Costantini et al., 2019; Danaher et al., 2014; Fried et al., 2018). In fact, when the tuning procedure selects a value of the corresponding tuning parameter equal or very close to zero, independent GGMs are estimated via typical graphical lasso. Therefore, JGL cannot hide differences nor inflate similarities across groups.

The JGL has been applied in the EDs research as well. Schlegl et al. (2021) used the FGL with *k*-fold cross-validation for parameter selection to estimate four different networks, one for each of the following groups: adolescents with AN, adults with AN, adolescents with BN, and adults with BN. Similarly, Smith et al. (2020) used the FGL to estimate and compare distinct networks from samples corresponding to either of the following groups: outpatients without ED diagnosis, outpatients with a lifetime attempt of suicide, and people with a current ED diagnosis. Martini et al. (2021) employed FGL to compare a sample of AN patients with a control group. Finally, (Forrest, Perkins, et al., 2019) jointly estimated and compared the network structures of two nonclinical samples of men with and without core ED symptoms.

#### Longitudinal and Personalized Networks

Time-series data allow for the estimation of personalized networks of two different kinds. *Temporal networks* incorporate consecutive temporal effects among symptoms by representing arrowhead edges pointing from one node to the other (or to itself in case of self-loops) when the first predicts the second in the next window of measurement. Edges are also weighted according to the regression parameters (Epskamp, van Borkulo, et al., 2018; Epskamp, Waldorp, et al., 2018). Temporal networks are commonly estimated as lag-1 Vector-Autoregression (VAR) models (van der Krieke et al., 2015), which consist of a set of regression equations on the given variables; these are all treated as endogenous, so they act as both outcome and predicted variables. The relations assessed through this method can also be interpreted in terms of Granger causality (Granger, 1969) by stating that, whenever an arrow from a time-varying symptom X to another time-varying symptom Y is found, then X *Granger-causes* Y, meaning that predictions of the value of Y based on its own past values and on the past values of X are better than predictions of Y based only on Y’s own past values. Clearly, Granger causality should not be erroneously understood as pure causality. Rather, Granger causality gives evidence of temporal prediction and thus it can be potentially indicative of causality in the sense that, although the existence of a causal relationship would imply the observation of a temporal prediction, the opposite is not true, since temporal links may also arise for other reasons. In addition, some temporal predictions can also be missed because of lack of statistical power or insufficient sized lag interval (Epskamp, van Borkulo, et al., 2018).

The residuals of the temporal VAR model are used to compute the so-called *contemporaneous network*, which can be estimated as a GGM model, with edges representing the partial correlation obtained after controlling for both temporal effects and all other variables in the same window of measurement (Epskamp, van Borkulo, et al., 2018). Together, the modeling framework including the estimation of both temporal and contemporaneous networks from a given dataset is called *graphical* VAR or GVAR (Wild et al., 2010). It can be computed with the help of the R package *graphicalVAR* (Epskamp, 2017), which allows for both regularized and unregularized estimations (Epskamp, Waldorp, et al., 2018).

When time-series of multiple subjects are available, it is possible to gain more insight into the network structure at the group-level by applying the *multilevel-VAR model*. For a given population, the average network parameters are called “fixed effects”, whereas the person-specific deviations from these fixed effects are called “random effects”. Random effects can be used to estimate intraindividual networks (temporal and contemporaneous) and to investigate interindividual differences (Bringmann et al., 2013). Fixed effects can be instead used to uncover information about average intraindividual effects. More generally, if a multivariate normal is assumed for all parameters, then estimating the GVAR model on longitudinal data allows for the decomposition of the variance into three different structures, namely temporal networks, contemporaneous networks and *between-subjects networks* (Epskamp, Waldorp, et al., 2018), where the latter is a GGM that allows for the examination of between-mean relationships for all individuals of the dataset. Estimation methods for multilevel-VAR models have been implemented in the *R* package *mlVAR* (Epskamp et al., 2017), which in particular permits to choose among different estimation procedures, such as sequential univariate multi-level estimation, multivariate Bayesian estimation, and fixed effects estimation.

In the context of the studies about EDs, Levinson et al. (2018b) first conducted a pilot study in which they collected longitudinal data from *N* = 66 participants by asking them to complete an EMA^4^ survey on ED cognitions and behaviors across one week. Using the *graphicalVAR* and *mlVAR* packages, they then estimated intraindividual networks to identify which symptoms maintain EDs within each individual as well as group-level networks, namely temporal, contemporaneous and between-subject networks. Later, they repeated an analogous study on a different dataset composed of longitudinal data of *N* = 1272 participants with the additional goal of comparing adolescents and adults network structures (Levinson et al., 2020). More recently, another study was conducted to exclusively estimate the individual networks of *N* = 34 participants with the aim of identifying and discussing a range of target symptoms for personalized ED treatment (Levinson et al., 2021).

### Approaches and Tools for Network Description

The analytical phase that follows the estimation of the network structure from data is designated to the investigation and interpretation of specific characteristics exhibited by either single nodes or groups of nodes jointly. Although a first visual inspection can give some general clues, many numerical methods can be employed to gain deeper insights.

#### Centrality Measures in GGMs

Depending on the type of network estimated, different measures can be used to investigate the role of each symptom in the network. In the case of undirected weighted networks, Opsahl et al. (2010) proposed a generalization to weighted networks (see Table 1) of the most common measures of node centrality that were originally designed by Freeman (1978) for binary networks, namely: degree, strength, closeness, and betweenness centrality.

**Table 1.** Definition and Interpretation of Common Centrality Measures. This table describes the centrality measures that are most commonly used in the analysis of undirected weighted networks, as formalized by Opsahl et al. (2010). The last row describes the (one-step) expected influence centrality (Robinaugh et al., 2016). For each measure, its definition, mathematical formulation and interpretation are given.

| Centrality Measure | Definition | Formulation | Interpretation |
| --- | --- | --- | --- |
| Degree | Number of edges connected to a node | $C_D(v) = \sum_{u \in \mathcal{V} \setminus \{v\}} e_{vu},$ where $e_{vu}$ is an edge between $u$ and $v$ | Higher values indicate higher centrality of that node in the network, that is, a central node is one with many connections. It is useful mainly in unweighted graphs. |
| Strength (S) | Sum of the absolute weights (e.g., partial correlation coefficients) of the edges connected to a node | $C_S(v) = \sum_{u \in \mathcal{V} \setminus \{v\}} \vert w_{uv}\vert ,$ where $w_{uv}$ is the absolute weight between $u$ and $v$ | It represents the likelihood that activation of a certain symptom will induce the activation of other direct symptoms (McNally, 2016). Just like the degree centrality, strength only accounts only for paths of unitary length. |
| Closeness (C) | Inverse of the sum of the (minimum) distance of a node to all the other nodes in the network. It can be normalized as the average length of the weighted shortest paths to all the other nodes | $C_C(v) = \left[ \sum_{u \in \mathcal{V} \setminus \{v\}} d(u, v) \right]^{-1}$ where $d(u, v)$ is the distance between $u$ and $v$ | It indicates the index of expected time until arrival of something flowing through the network (Borgatti, 2005). In simple words, a node with high closeness is one that is close, on average, to other nodes. Hence, nodes with low closeness centrality are likely to be sooner influenced by changes in the network. |
| Betweenness (B) | Number of the geodesics between any two other nodes | $C_B(v) = \frac{g_{uz}(v)}{g_{uz}},$ | This measure plays an important role in the assessment of comorbidities, since symptoms |
| | that pass through a given node | where $g_{uz}$ is the number of binary shortest paths between two nodes, and $g_{uz}(v)$ is the number of those paths that go through node $v$ | with high betweenness centrality usually serve as bridge symptoms; when activated, they are likely to spread to both syndromic clusters (McNally, 2016). In other words, a high-betweenness node is one that acts as a bridge in the communication with other nodes. |
| (One-Step) Expected Influence (EI) | It computes the strength of a node by considering both positive and negative correlations | $EI(v) = \sum_{u \in V \setminus \{v\}} w_{uv},$ where $w_{uv}$ is an edge between $u$ and $v$ | It exhibits the same performance of strength when the network only contains positive weights, whereas it outperforms strength as the number of negative weights increases. It has been proven that EI is a better predictor of declines in the severity of symptoms over time. It is particularly useful when the aim is to identify target symptoms for therapeutic deactivation since these can only be identified through negative correlations. |

However, all the measures above only consider weights in absolute value with the consequence that two nodes may have same centrality but opposing effects on the rest of the network. For example, an increase in Node A can cause an increase in Node B (positive influence) with the same strength that an increase in Node C causes a decrease in Node D (negative influence). In this case, Node A and C will have the same centrality index but an opposite effect. Moreover, a node with a similar number of strong positive and negative edges may have little to no overall network activation impact, that is, it might be highly central without being highly influential. Hence, a new centrality metric called *expected influence* (EI) has been introduced to consider both negative and positive edges (see Table 1, last row; Robinaugh et al., 2016).

As argued by Bringmann et al. (2019), betweenness and closeness centrality do not seem to be especially suitable as measures of node importance. Hence, results about these metrics should be interpreted with care. As an additional proof about this fact, many articles reported betweenness and closeness centrality as not satisfying the minimum stability results and did not include their estimation in the network description (Aloi et al., 2021; Calugi et al., 2021; Forrest et al., 2018; Monteleone et al., 2020; Rodgers et al., 2018; Smith et al., 2019; Vanzhula et al., 2019). Others decided instead to take these measures into account but did not assess their stability (Forbush et al., 2016; Olatunji et al., 2018).

Throughout the large number of studies reviewed, few symptoms appeared among the most central ones across heterogeneous samples and estimation techniques (see Table 2).

**Table 2.** Central Symptoms in Psychopathology Networks. This table summarizes the symptoms (left column) that were most often identified as central in the psychopathology networks built from psychometric data concerning either specific or nonspecific ED symptoms. The central column indicates the metric used (S = strength, EI = expected influence, C = closeness, and B = betweenness). The column on the right contains the list of papers that find the symptom as a result of their analysis.

| Symptom | Centr. Measure | References |
| --- | --- | --- |
| Shape and weight preoccupation and overvaluation, body dissatisfaction | S and EI | Calugi et al., 2020; DuBois et al., 2017; Elliott et al., 2020; Forbush et al., 2016; Forrest et al., 2018; Forrest, Perkins, et al., 2019; Forrest, Sarfan, et al., 2019; Goldschmidt et al., 2018; Kenny et al., 2021; Kinkel-Ram et al., 2021; Levinson et al., 2021; Levinson et al., 2020; Mares et al., 2021; Meier et al., 2020; Rodgers et al., 2019; Vanzhula et al., 2019; 2021; Wang et al., 2019 |
| Fear of weight gain | S and EI | Calugi et al., 2021; Calugi et al., 2020; Elliott et al., 2020; Forrest et al., 2018; Goldschmidt et al., 2018; Hagan et al., 2021; Kinkel-Ram et al., 2021; Levinson et al., 2021; Levinson et al., 2020; Levinson et al., 2017; Meier et al., 2020; Perez et al., 2021; Rodgers et al., 2019; Vanzhula et al., 2019 |
| Drive for thinness | S, EI, C and B | Brown et al., 2020; Cascino et al., 2019; Christian et al., 2021; De Paoli et al., 2020; Elliott et al., 2020; Forrest, Perkins, et al., 2019; Hagan et al., 2021; Kinkel-Ram et al., 2021; Levinson et al., 2021; Levinson et al., 2018b; Monteleone, Mereu, et al., 2019; Olatunji et al., 2018; Perez et al., 2021; Rodgers et al., 2018; Sahlan, Williams, et al., 2021; Schlegl et al., 2021; Smith et al., 2019; Solmi et al., 2019; Vanzhula et al., 2019 |
| Ineffectiveness (i.e., feeling of inadequacy, insecurity, worthlessness and having no control over one's life) | S, EI, C and B | de Vos et al., 2021; Martini et al., 2021; Monteleone, Cascino, et al., 2019; Monteleone, Tzischinsky, et al., 2022; Olatunji et al., 2018; Ralph-Nearman et al., 2021; Sahlan, Williams, et al., 2021; Schlegl et al., 2021; Solmi et al., 2018; Solmi et al., 2019; Vervaet et al., 2021 |
| Lack of interoceptive awareness (i.e., ability to identify, access, understand, and respond properly to the patterns of internal signal) | S, C, and B | Cascino et al., 2019; De Paoli et al., 2020; Monteleone, Cascino, et al., 2019; Monteleone, Tzischinsky, et al., 2022; Olatunji et al., 2018; Solmi et al., 2018; Solmi et al., 2019; Vervaet et al., 2021 |
| Social insecurity, in particular avoidance of social eating | S, EI, C and B | Bronstein et al., 2022; Cascino et al., 2019; Kerr-Gaffney et al., 2020; Martini et al., 2021; Olatunji et al., 2018; Ralph-Nearman et al., 2021; Wang et al., 2019 |
| Dietary restriction | S and EI | Calugi et al., 2021; Calugi et al., 2020; Forrest, Sarfan, et al., 2019; Goldschmidt et al., 2018; Hagan et al., 2021; Mares et al., 2021; Monteleone et al., 2020; Rodgers et al., 2019 |
| Depressed mood | S and EI | Beauchamp et al., 2021; Bronstein et al., 2022; de Vos et al., 2021; Rodgers et al., 2019; Solmi et al., 2018; Solmi et al., 2019 |
| Binge-eating | S | Christian et al., 2020; 2021; Levinson, et al., 2018a; Levinson et al., 2018b; Levinson et al., 2020; Perko et al., 2019; Vanzhula et al., 2019 |

Diverging from the other studies, 3 out of 56 papers also assessed the importance of nodes using *key players* analysis. Specifically, they identified the nodes that, when removed, would result in a maximally disconnected residual network. In other words, a treatment targeting these key nodes is expected to slow the cascade of symptoms through the ED network. Analyzing the symptom network of a sample of participants with mixed ED diagnoses, Forbush et al. (2016) identified its key players to be: *people encouraged me to eat more, need to exercise nearly every day,* and *try to avoid foods with high calorie content.* Perko et al. (2019) employed this metric to assess sex differences in ED symptoms, but they found that the key players were, in both cases, items related restricting dieting and binge eating. More recently, Liebman et al. (2021) explored the associations between posttraumatic stress disorder and ED symptom in presence of at least one experience of childhood abuse and found that the key players of the network were: *purging, negative alterations in cognitions and mood,* and *hyperarousal*.

#### Interpretation and Measures of Importance in Bayesian Networks

When interpreting a Bayesian network visualized as a DAG, node importance cannot be evaluated through the above-mentioned centrality measures for undirected graphs. Rather, Rodgers et al. (2019) proposed to assess this property based on the relative contribution of each symptom to the overall model fit of the Bayesian network. In particular, they evaluated the *scaled contribution of each symptom to BIC* for three different DAGs: one estimated from the full sample, one from the subsample of participants who experience childhood abuse in addition to having received an ED diagnosis, and one for the subsample of those that only suffer from an ED. The result was that in the first and third group the most important symptoms were *overvaluation of shape and weight, depressed mood* and *eating large amounts of food*, whereas in the second group a different pattern of relationships among symptoms emerged, with *depressed mood, dietary restraint, self-induced vomiting,* and *driven exercise* being the most important driving symptoms of the disorder. The dissimilarity found was consistent with the concept of *maltreated ecophenotype* (Teicher & Samson, 2013), according to which distinct subtypes of a given disorder may be developed as a consequence of abuse and trauma. This hypothesis has been also confirmed by other experimental studies on different psychiatric disorders including EDs (Monteleone, Cascino, et al., 2021).

#### Understanding Comorbidities Between EDs and Other Psychopathologies

With the specific aim of identifying bridge symptoms, four network statistics have been developed and implemented in the *R* package *networktools*; importantly, since they are not specific to the type of network estimated, they can be readily applied to intraindividual networks, group-level networks, and other networks different from the psychopathology ones (Jones et al., 2021).

They are defined as follows:

● *Bridge strength* (BS), that is, the sum of the absolute value of every edge which connects a given symptom to symptoms belonging to other disorders. *Bridge in-strength* and *bridge out-strength* can be analogously defined in directed networks by considering only the subset of edges that are, respectively, directed toward the node or issued from a node.
● *Bridge expected influence* (BEI), which is defined just like the bridge strength but without taking the absolute value. In directed networks, only edges issuing from a node are summed.
● *Bridge Closeness* (BC), that is, the average distance from a given node to all nodes outside of its own disorder.
● *Bridge betweenness* (BB), that is, the number of times a given node lies on the shortest path between any two nodes belonging to two distinct disorders (including the one it belongs to).

Refer to Table 3 for the list of the bridge symptoms between EDs and other psychopathologies that have been assessed through the metrics defined above in some of the papers under review.

**Table 3.**
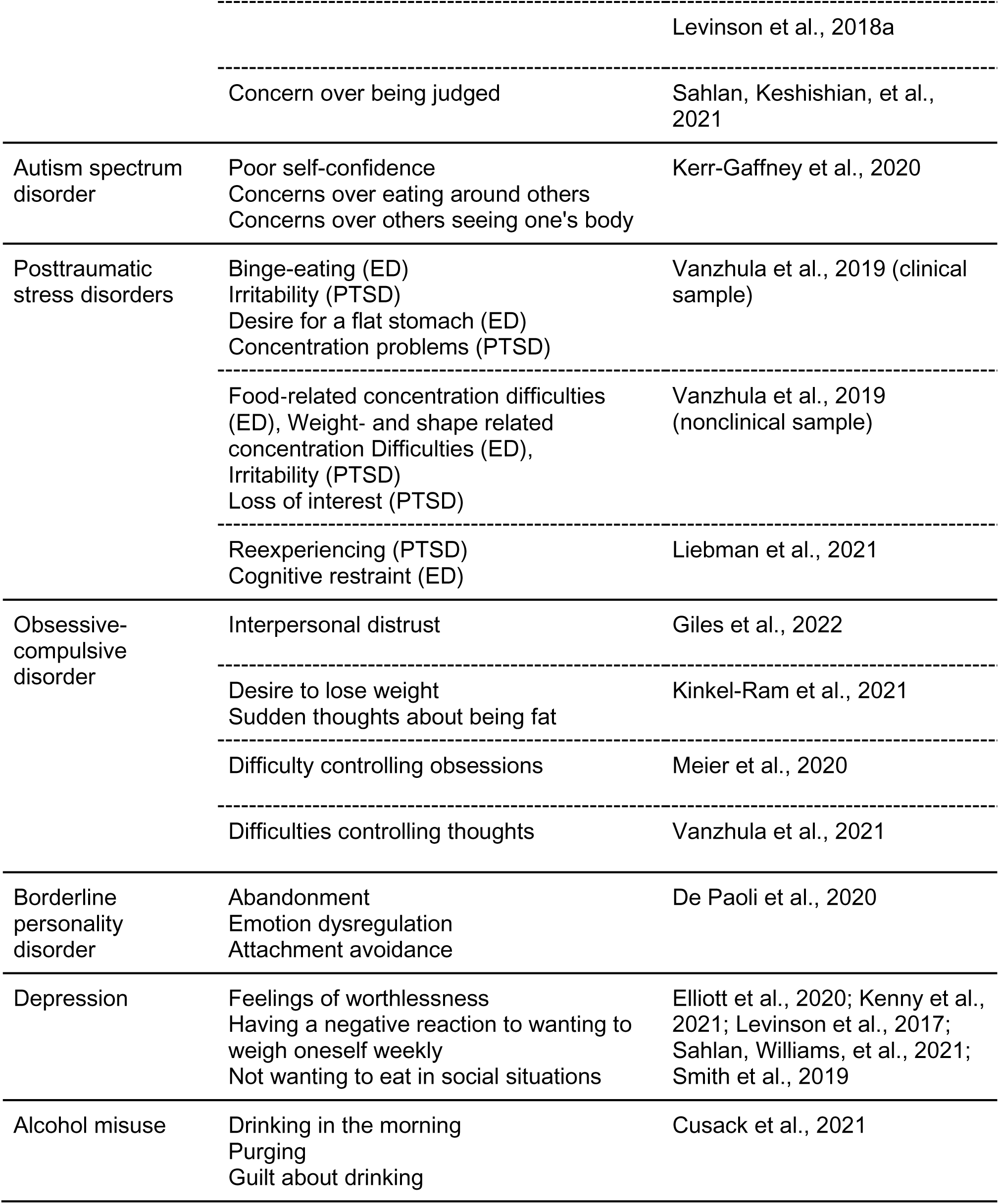
Bridge Symptoms Between Eating Disorders and Other Psychopathologies. This table lists, for each of the analyzed comorbidity with EDs (left column), the symptoms with highest bridge EI (center column) as reported in the reference papers (right column).

#### Assessing the Role of the External Field in the Development and Maintenance of EDs

Interactions between ED specific symptoms and various elements of the external field have also been investigated. Few studies used the bridge centrality measures to accomplish this task. Monteleone et al. (2019; 2021) chose instead to adopt a different approach to explore the psychological pathways through which childhood maltreatment (CM) experiences promote the development of ED core symptoms. Namely, they first selected few variables from items and scores of different psychometric questionnaires in order to build symptom networks for each ED diagnosis, then the shortest path between any CM and ED node was computed using Dijkstra’s algorithm, and finally they used mediation analysis to confirm the mediation role of the symptoms included in the shortest pathways from CM to ED specific symptoms. All the other studies considered all variables as a single community and chose to determine the core symptoms using the centrality measures in their classical form with the specific aim of verifying whether EDs are mainly maintained by ED specific symptoms or rather by nonspecific ones. The results achieved throughout the papers under review are summarized in Table 4.

**Table 4.**
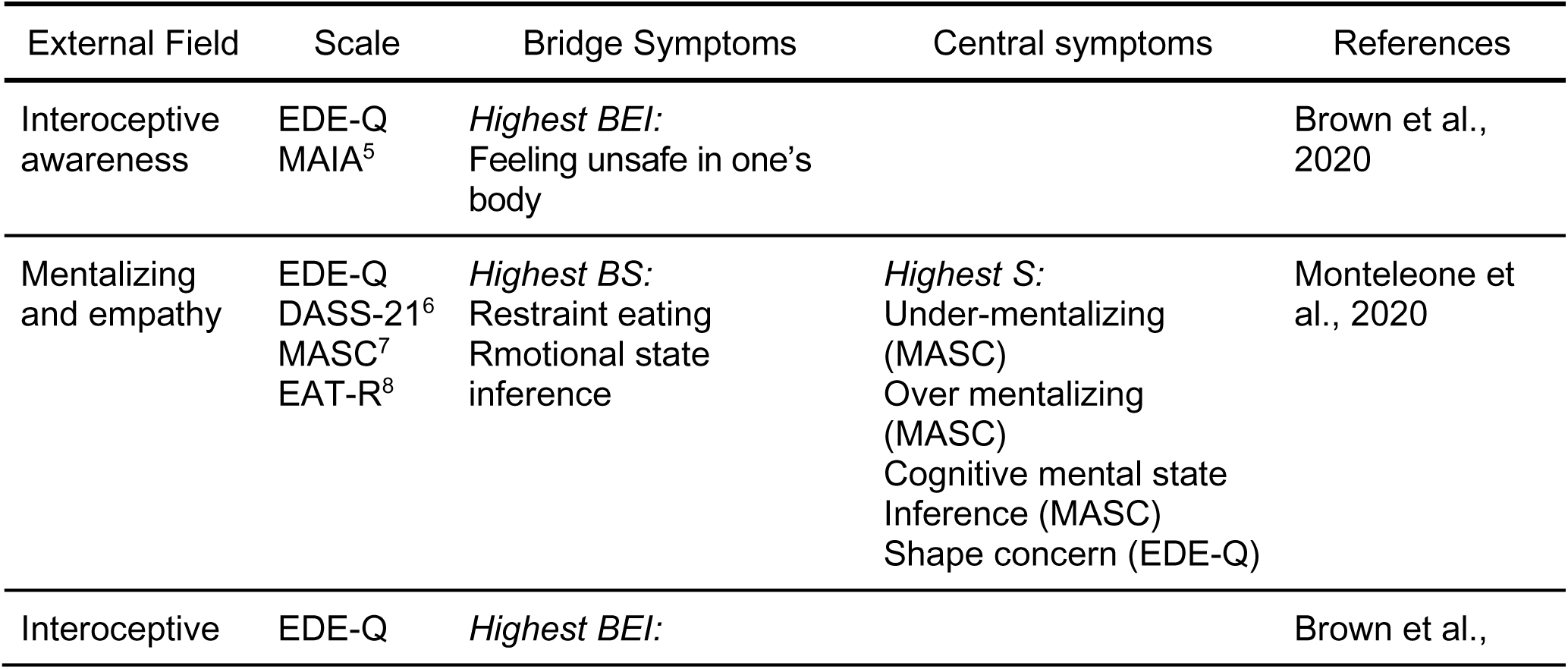
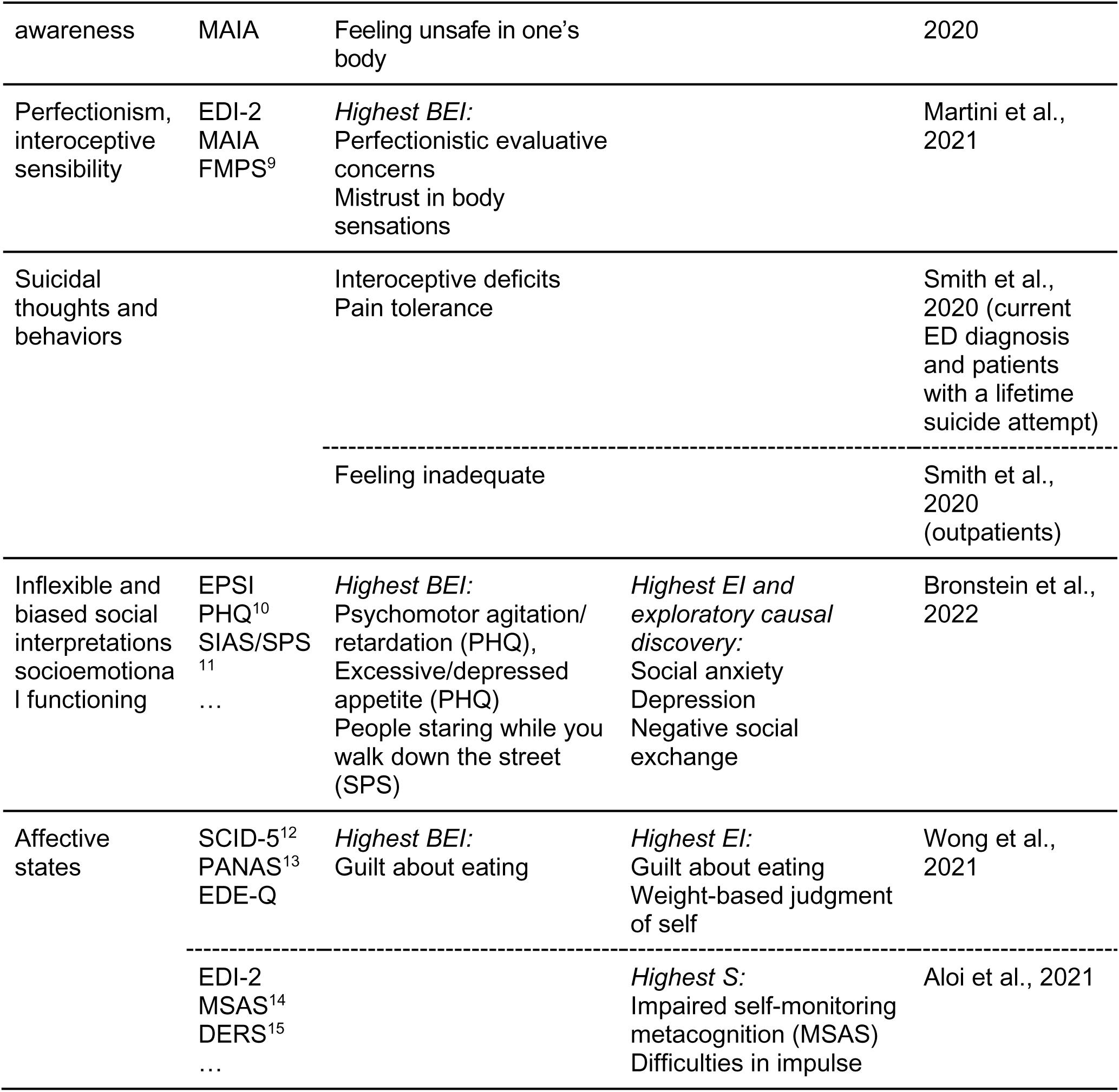
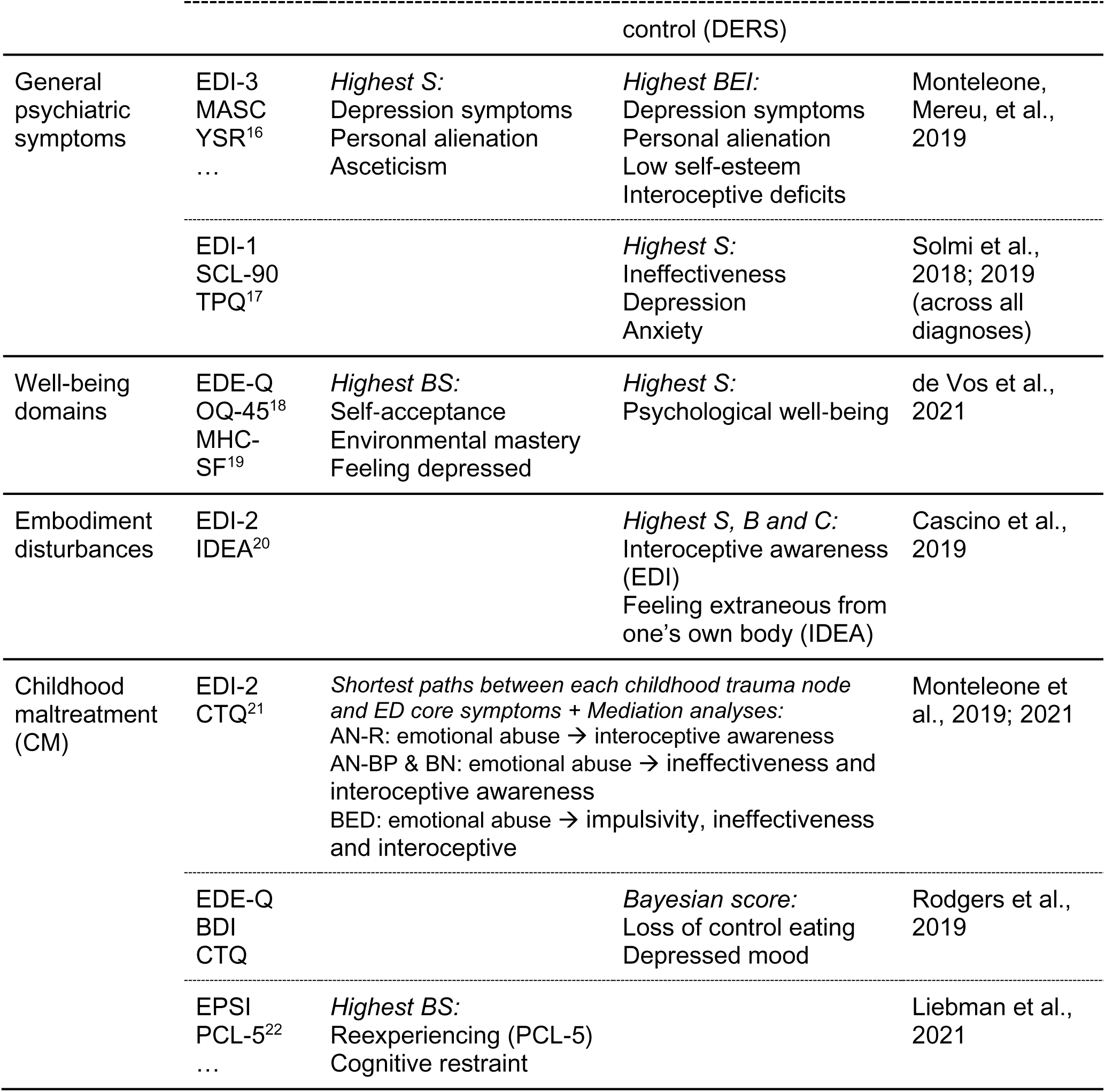
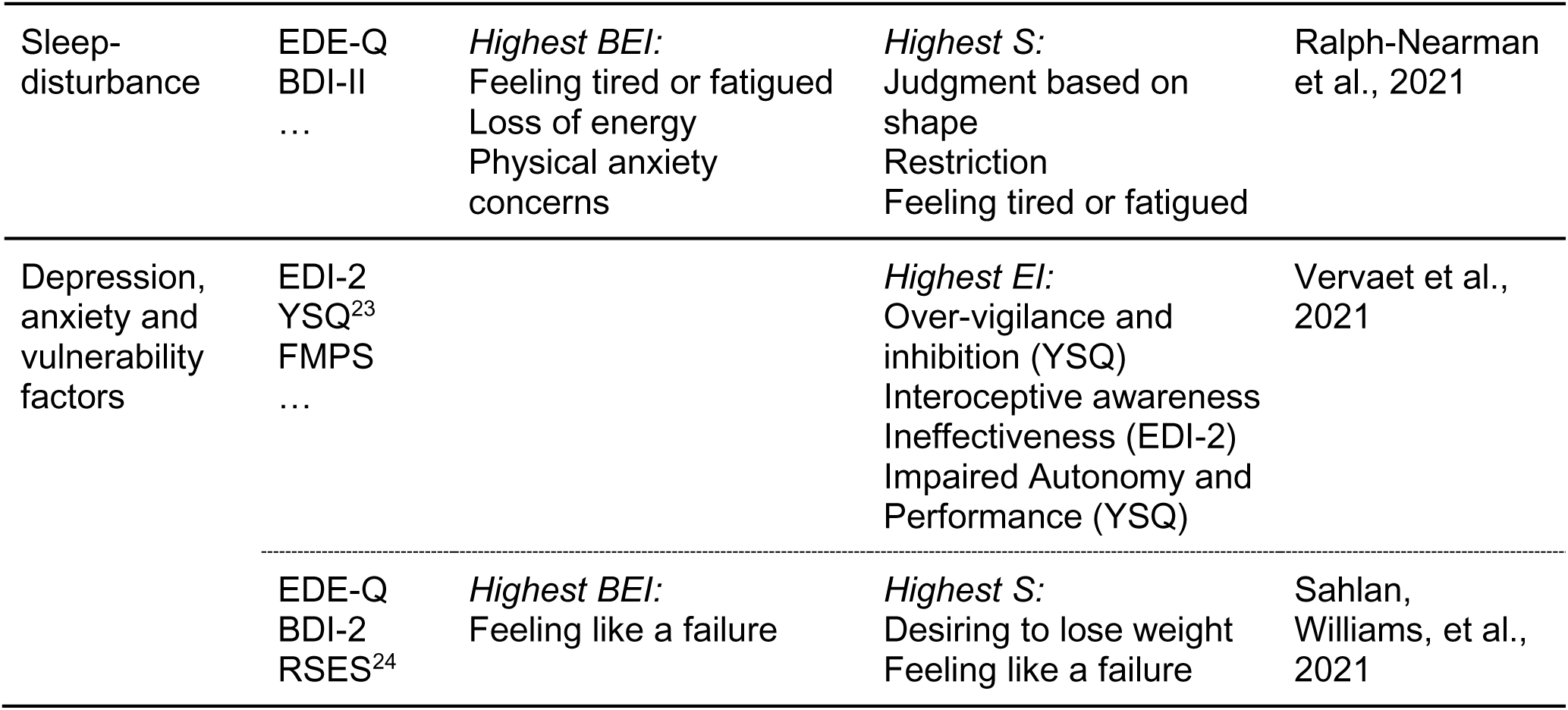
Interaction Between Eating Disorders and the External Field. This table summarizes the results achieved throughout the papers under review concerning the role of the external field in the development and maintenance of EDs. For each specific external factor, many details are given, namely: the psychometric assessment tool used to assess it, both bridge and central symptoms identified in the psychopathology network, and the reference paper.

#### Longitudinal Studies

All longitudinal studies that used the *graphicalVAR* and *mlVAR* for network estimation also computed node centrality through the following measures: strength for between-subject and contemporaneous networks, in-strength and out-strength for temporal networks. In the latter case, that specific choice of metrics was taken to underlie the different impact of incoming against outgoing edges. More precisely, *In-strength* is given by the sum of links pointing towards the node and indicates how much information that node is receiving from directly connected nodes. Instead, *out-strength* is given by the sum of links pointing from one node to all the other and indicates how much information that node is sending to directly connected nodes. This distinction has the precious advantage of suggesting which nodes (i.e., those with highest out-strength), have the potentiality of having downstream effects on other symptoms if treated (Levinson et al., 2021).

As for the group-level temporal networks, the symptoms with highest in-strength centrality were *desire to be thin, body checking* (Levinson et al., 2018b), *fasting, fear of weight gain* and *feeling fat* (Levinson et al., 2021; Levinson et al., 2020). Those with highest out-strength centrality were *exercise, binge eating* (Levinson et al., 2018b), *feeling fat* and *fear of weight gain* (Levinson et al., 2020). The strongest symptoms in the contemporaneous and between-subject group-level networks were *desire to be thin* (Levinson et al., 2018b), and *feeling fat* (Levinson et al., 2020). Among the individual networks, nodes with highest in-strength centrality in the temporal networks were *overvaluation of weight and shape, fear of weight gain,* and *body dissatisfaction* (Levinson et al., 2020); while those with highest out-strength were: *exercise, fear of weight gain, overvaluation of weight and shape (Levinson et al., 2020)*, and *body dissatisfaction* (Levinson et al., 2021). Finally, strongest symptoms in the individual contemporaneous networks were *thinking about dieting, binge eating* (Levinson et al., 2018b), *fear of weight gain (Levinson et al., 2020)*, *body dissatisfaction* and *drive for thinness* (Levinson et al., 2021)

#### Pre- to Post-treatment Studies

Few studies have been conducted to compare pre- and post-treatment network models. In particular, they all estimate and compare GGM networks at each time point (mainly at admission and discharge, in few cases also at follow-up) and few studies also assessed the prognostic value of the most central nodes at baseline through linear or logistic regression. One of them (Elliott et al., 2020) first computed zero-order correlations between each symptom at baseline and three outcome measures (i.e., treatment recovery status, clinical impairment, and posttreatment BMI) and then tested whether these prognostic values were associated with the expected influence of symptoms at baseline via linear regression. The results of this study show that EI values remained constant across all time points, with the strongest nodes being *feeling fat, fear of weight gain, discomfort seeing one’s own body, dissatisfaction with weight,* and *a strong desire to lose weight*. The authors also found that more severe symptom levels were associated with a lower possibility of recovery, higher clinical impairment and higher BMI. Finally, they observed that centrality of symptoms at baseline was significantly associated with prognostic values for both recovery status and clinical impairment. Similarly, Hagan et al. (2021) found that pretreatment central symptoms in adolescents with AN significantly predicted early response but did not predict remission. A third study (Olatunji et al., 2018) also found that the most central symptoms, namely *interoceptive awareness* and *ineffectiveness*, did not change after treatment. The authors also used multiple regression to test whether the identified core symptoms predicted outcomes (BMI, depression, and anxiety) at discharge. Their hypothesis was confirmed for BMI and depression but not for anxiety. Finally, Brown et al. (2020) showed that stronger *desire to lose weight* at admission was associated with lower likelihood of achieving remission at discharge.

Many other studies only estimated and compared the centrality of symptoms at different time points. A change in the role of certain symptoms was found, with the strongest at baseline being *fearing weight gain, dietary rules* (Calugi et al., 2021), *eating disorder-related impairment, self-esteem* and *shape concern* (Hilbert et al., 2020), and the strongest at discharge being *dietary restraint* (Calugi et al., 2021; Hilbert et al., 2020), *food preoccupation, feelings of fatness* and *discomfort seeing its own body* (Calugi et al., 2021). Smith et al. (2019) only computed centrality measures of the admission network, finding that the strongest symptoms were: *shape and weight-related concentration difficulties, general concentration difficulties, guilt about eating, desire to lose weight,* and *nervousness*.

### Network Stability Analysis

Network stability analysis has been conducted in most of the papers under review, especially in those published after 2018, when Epskamp and his colleagues Borsboom and Fried (2018) proposed precise guidelines for this task. In particular, they suggested specific methods to assess the robustness of the model at three distinct levels: accuracy of the estimated edge weights, stability of the order of centrality indices, and difference between specific edge weights or centrality indices.

#### Accuracy of edge weights

The accuracy of edge weights can be evaluated by constructing intervals that reflect the sensitivity of edge weight estimates to sampling error, such as confidence intervals (CIs), credibility intervals and bootstrapped intervals (Borsboom et al., 2021). When handling ordinal data as in the current study, it has been suggested to derive the (1-α) CIs via nonparametric bootstrap at a given confidence level, for example, α = 0.05 (Epskamp, Borsboom, et al., 2018). The narrower the CIs, the more likely is that the estimated edge weight is close to its real value, since in 95% of the cases such a CI will contain the true value of the parameter. Although large CIs can result in a poor accuracy for centrality indices, they do not influence the presence of an edge, nor its sign, as these properties are already assessed by LASSO. Moreover, it should be noticed that since we use regularization to estimate the network structure, all edge weight estimates are biased towards zero and, consequently, all sampling distributions are biased towards zero as well, just like CIs are not centered around the true unbiased parameter value anymore. This implies that, when interpreting the quantiles of the bootstrapped sampling distribution, if they overlap with zero it could be that the corresponding CI does not overlap with zero, while if they do not overlap with zero, then also the corresponding CI does not overlap with zero (Fried et al., 2020). In other words, CIs should not be interpreted as significance tests to zero, but only to show the accuracy of edge weight estimates by evaluating the size of CIs and to compare edges to one another by checking if the corresponding CIs do not overlap; if so, then we can conclude that they significantly differ at the given significance level, in the other case, then we cannot infer the contrary since they might still significantly differ (Epskamp, Borsboom, et al., 2018).

#### Stability of centrality indices

As noticed by Epskamp and colleagues (2018), the same bootstrap technique cannot be used to construct true CIs around the centrality indices. As an alternative, they proposed to investigate the stability of the order of centrality indices based on subsets of the data, that is, by comparing the order of centrality indices after re-estimating the network with fewer cases or nodes. When this is done for various proportions of cases to drop, then one can also assess the correlation between the original centrality indices and those obtained from subsets. If this correlation keeps stable after dropping a considerable proportion of the cases (e.g., 10%), then the interpretations of centralities can be considered stable. This method has been called *case-dropping subset bootstrap*. Exploiting this technique, a quantification of the stability of a centrality index can be given by the *correlation stability coefficient* (CS-coefficient), a measure representing the maximum proportion of cases that can be dropped, such that the correlation between original centrality indices and that of networks built on subsets with 95 % probability is still equal or higher than a given value which is set to 0.7 by default. As a cutoff score for interpreting an estimated centrality index as stable, Epskamp and colleagues (2018) suggested that the CS-coefficient value should be above 0.5 or in any case not below 0.25. As already mentioned, this cutoff was not reached by the CS-coefficient of closeness and betweenness centrality in almost all cases. On the other side, strength and expected influence always attained pretty good values, usually significantly above the threshold.

#### Methodological differences in edge weights and centralities

The last technique proposed by Epskamp (2018) is the *bootstrapped difference test*, which is a null-hypothesis test used to assess whether the edge weights or centralities differ from one another. This is accomplished by considering the difference between the two bootstrap values of edge weights or centrality indices under study and constructing a bootstrapped CI around those difference scores. If zero is not in the bootstrapped CI, then the null hypothesis is rejected, meaning that there is evidence that two values differ from one-another. On the other hand, it should be noticed that not rejecting the null-hypothesis is not sufficient for inferring that the null-hypothesis is true (2018). Finally, Epskamp et al. (2018) emphasized and warned about the fact that this technique does not take into account any correction for multiple testing, since applying Bonferroni correction is not feasible in practice in this context. As a consequence, as the number of performed significance tests increases, the probability of finding several significant results purely by chance (Type 1 error) also increases.

#### Other approaches for stability estimation

The above-mentioned stability techniques do not apply to Bayesian networks. As an alternative, Rodgers et al. (2019) quantified the arc strength, that is, the degree of confidence it is possible to have when interpreting specific pathways, through a bootstrapping procedure introduced by Friedman et al. (2013; 1999) and implemented in the *bnlearn R* package (Scutari, 2009). The general idea behind this procedure is that we should be more confident on features that would still be induced when we perturb the data. Therefore, in a nonparametric bootstrap setting, one first generates perturbations by re-sampling with replacement from the given dataset and then examines how many of the perturbed structures exhibit the feature under study that, in this specific case, corresponds to the presence and direction of each edge. Their relative frequency across the bootstrapped samples gives an estimation of each arc strength (Scutari & Nagarajan, 2013). Rodgers et al. (2019) reported an average frequency of 84% concerning the presence of edges correctly identified across bootstrapped samples, and an average frequency of 63% for their direction. The arc *fear of gaining weight → cognitive restraint* had the highest presence frequency of 99%, while the arc *preoccupation with eating and body image → depressed mood* had the highest direction frequency of 88%.

### Common Methods for Network Comparison

Many of the studies under review aimed at comparing network structures across different populations. A specific tool to accomplish this task, namely the *Network Comparison Test* (NCT; van Borkulo et al., 2015) has been devised and implemented in the *R* package *NetworkComparisonTest* (Van Borkulo et al., 2017). Compared to other statistical tests, the NCT overcomes the usual assumption of normality and possible improper null hypothesis that are unsuitable for the regularized parameters that results after GGM estimation with LASSO regularization, the most typical method employed in the network approach to psychopathology.

The NCT is a 2-tailed permutation test in which the difference between two groups is calculated repeatedly for randomly regrouped individuals. It consists of three steps: first, the network structure is estimated for both groups using the original data and the metric of interest is calculated; second, data is permuted iteratively to rearrange group memberships, networks are then re-estimated, and metrics are calculated based on permuted data to create a reference distribution; finally, the significance of the observed test statistic is evaluated by comparing it to the reference distribution. In particular, the *p*-value equals the proportion of test statistics that are at least as extreme as the observed test statistic. Thus, the null hypothesis that the two networks under comparison are the same can be rejected if the latter is larger than expected (i.e., *p*-value < 0.05).

As for the test statistics that can be used to compare networks, van Borkulo et al. (2017) proposed three metrics that represent both global and local differences, namely invariance of network structure and global strength for the global differences, and invariance of edge strength for the local differences (see Table 5).

**Table 5.**
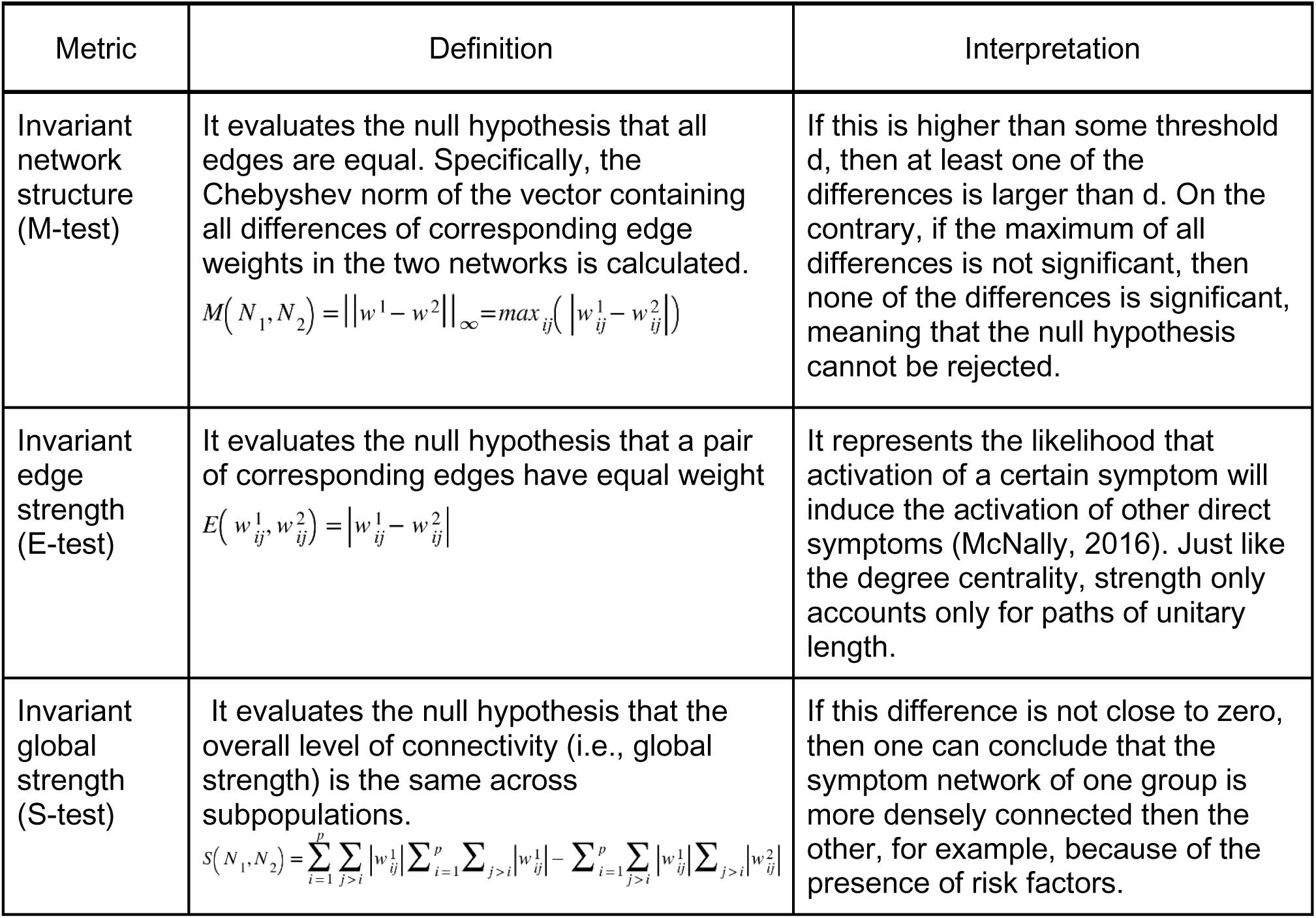
Network Comparison. Definition and interpretation of the three test statistics introduced by van Borkulo et al. (2017) to compare a pair of networks based on their global and local differences

The above test statistics have already been used in several studies about EDs. Significant results concerning the invariant network structure have been found in various comparisons, in particular: clinical versus nonclinical subsamples of networks assessing comorbidity between EDs and different features of social anxiety disorder (Levinson et al., 2018a); admission against discharge network (Calugi et al., 2021); adolescents versus adults with AN and BN, with the exception of the comparison between adolescents with BN and adults with BN (Schlegl et al., 2021).

Moreover, significant results have been obtained when computing the invariant global strength for many pairs of networks, among those: groups of individuals with low versus high levels of overvaluation of shape and weight, with the latter resulting in higher connectivity (DuBois et al., 2017); clinical versus nonclinical samples, with a higher density in the former (Vanzhula et al., 2019); groups of individuals split based on the median value of the EDE-Q global scores at admission and discharge, with denser networks at admission predicting less change in ED symptomatology during treatment (Smith et al., 2019); pre-to posttreatment networks, with increased connectivity in the latter (Hilbert et al., 2020); admission against discharge network, with decreased connectivity in the latter (Calugi et al., 2021); adolescents with AN versus adolescents with BN, adolescents with AN and adults with BN, both cases with higher global strength in the AN sample (Schlegl et al., 2021); men with core ED symptoms versus men without them (Forrest, Perkins, et al., 2019); men versus women (Perko et al., 2019); across developmental stages (Christian et al., 2020);

Finally, having found a significant variation in the network structures, Calugi et al. (2021) also tested the change in weight of links from admission to discharge. He identified few connections that were stronger at baseline than at discharge, namely *feelings of fatness* and *desiring weight loss*, *BMI* and *vomiting to control shape or weight*, and others whose relationship grew at discharge, namely *food preoccupation* and *desiring weight loss*, *fear of losing control overeating* and *vomiting to control shape or weight*, *feelings of fatness* and *dissatisfaction with weight and shape*. Schlegl et al. (2021) reported instead the percentage of edges that were significantly different in each pair of networks which resulted to have significant invariant network structure statistics. The values found ranged from 2.56% for the adolescents with AN versus adults with AN comparison to 10% for the adolescents with AN versus adults with BN comparison.

Remarkably, many studies reported no differences in network structure nor in global strength with regards to different ED diagnoses (de Vos et al., 2021; Goldschmidt et al., 2018; Mares et al., 2021), age (Brown et al., 2020; Calugi et al., 2020; Sahlan, Williams, et al., 2021), and sex (Sahlan, Keshishian, et al., 2021; Sahlan, Williams, et al., 2021).

One study assessing the network differences from admission to discharge, although not finding any significant changes in the global strength, reported a significant effect of time on symptom severity, indicating decreases in ED, depression, and anxiety symptoms, with medium to large effect sizes (Smith et al., 2019). These results were assessed through repeated measures multivariate analysis of variance (RM MANOVA), which is a statistical technique to determine the degree to which multiple dependent variables (e.g., total scores of psychometric assessment tests) vary across time points.

## Discussion

Throughout this work, we first recalled the recent and promising field of psychometric network analysis, and we then outlined a comprehensive review of its methods applied to the study of eating disorders. What emerged from our data is a coherent image describing strong symptom interconnections between specific and nonspecific ED symptoms, as well as important roles in the arising and maintaining of ED shared by both types of symptoms. This points in the clear direction that clinical intervention should also address general psychological distresses, such as ineffectiveness and interoceptive awareness, instead of ED specific symptoms only. However, this image is still incomplete, firstly because the results already obtained have not received an experimental verification yet. Moreover, the results of the present systematic review suggest that methodologies capable of integrating different input information into one single network structure might be essential to uncover the dynamic of EDs, but, to our knowledge, they still need to be implemented and verified in a clinical setting. A possible direction to accomplish this task might be the multilayer network approach (Kivelä et al., 2014), according to which a complex system can be modeled as a network of networks, in other words, as a set of multiple layers with connections between and within them (de Boer et al. 2021). In the case of mental disorders, one can think of extending the study of symptom networks with other entities (such as genetic factors, brain structure and functional connectivity, environmental factors) as layers in a multilayer network. An attempt to implement this approach has already been proposed to integrate multiple levels of personality, namely neural and psychological constituents (Brooks et al. 2020), but an application to psychopathologies is also advised (Braun et al., 2018).

Most of the reviewed studies were based on cross-sectional data retrieved from structured psychometric questionnaires administered to subpopulations of individuals diagnosed with an ED disorder. The most widely used questionnaires to assess ED specific symptoms were EDE-Q and EDI-II, which also accounts for general psychological factors. Other questionnaires widely used to assess nonspecific ED symptoms were SCL-90 and BDI.

Only in a few cases, mostly conducted by Levinson and colleagues (2021; 2018; 2020), the analysis was carried out on panel data collected by means of EMA methods or repeated administration of one or more specific questionnaires to the same sample at different time points. These were the only studies that were able to answer research questions about the dynamic of symptom networks and the intraindividual network structure. Due to the limited number of publications and the considerable clinical implications, our data suggest that future research should give this issue more focus.

With regard to the general characteristics of the participants, the most blatant peculiarity is surely the clear prevalence of female patients. Only one study (Forrest, Perkins, et al., 2019) reported a greater percentage of male participants that, however, were not recruited in a clinical setting.

Almost all studies included in their analysis a node selection step to eliminate redundant items and obtain more accurate results. Those relying on cross-sectional data mainly used a Gaussian Graphical Model with LASSO regularization technique to estimate an undirected symptom network. Only one study (Forbush et al., 2016) was found to employ nonregularized methods, such as association and concentration graphs, and only one (Rodgers et al., 2019) produced a directed Bayesian network. The parallel estimation of symptom networks on different subsamples was achieved in few cases through the FGL technique (Forrest, Perkins, et al., 2019; Martini et al., 2021; Schlegl et al., 2021; Smith et al., 2020). Finally, different studies based on panel data were also found to employ mlVAR to estimate between-subject networks and graphicalVAR to estimate temporal and contemporaneous networks (Levinson et al., 2021; Levinson et al., 2018; Levinson et al., 2020).

The network description step was focused on the identification of the core symptoms and of the bridge symptoms in case of research questions concerning comorbidities. As for the first point, our data suggest that both specific and nonspecific ED symptoms are central for the development and maintenance of ED psychopathology, in particular *shape and weight overvaluation, body dissatisfaction, fear of weight gain, drive for thinness, ineffectiveness, lack of interoceptive awareness,* and *social insecurity.* As for the second point, *avoidance of social eating* and *lack of self-confidence* were found to bridge ED with anxiety disorders, whereas *feelings of worthlessness, having a negative reaction to wanting to weigh oneself weekly* and *not wanting to eat in social situations* were found to bridge ED with depression. When exploring the relationship with the external field, *emotional abuse during childhood* has been identified as a highly influential variable for the development of any ED (Monteleone, Cascino, et al., 2019; Monteleone, Tzischinsky, et al., 2022). In all longitudinal studies ED specific symptoms like *overvaluation of weight and shape* and *fear of weight gain* reported the highest in-strength and out-strength centrality. Finally, the pre- to posttreatment comparison revealed that central symptoms remained constant across all time points, with more severe symptom levels associated with a lower possibility of recovery, higher clinical impairment (Elliott et al., 2020; Olatunji et al., 2018). A change in the role of certain symptoms was found instead by Hilbert et al. (2020) and Calugi et al. (2021).

Network stability analysis has been conducted in almost all papers under review (explicitly reported in 49 out of 56). In particular, almost all authors employed the bootstrap methods previously described to compute the accuracy of the estimated edge weights, the stability of the order of centrality indices, and difference between specific edge weights or centrality indices. One common finding is that strength and expected influence generally performs much better, i.e., are more stable, than closeness and betweenness centrality and can thus be assumed to be more reliable indices of the centrality of symptoms.

The NCT was employed to reveal differences in the symptom networks of samples with different characteristics. In particular, our study reported many cases in which similarities in network structures were found, although with different levels of connectivity. An important note should be mentioned here about the global strength of pre- and posttreatment networks. According to the network theory of psychopathology, effective treatment should lead to a decrease in network connectivity and its self-sustaining character, but this assumption was not met in some studies (Hilbert et al., 2020), suggesting that further research on the predictive value of network variables in the therapeutic outcome is needed.

As a final remark, our data highlight the fact that the state-of-the-art procedures are all based on a collection of *R* packages specifically designed for the network analysis of psychometric data. To our knowledge, no study employed other software commonly used in network science, such as Cytoscape (Shannon et al., 2003), Pajek (Batagelj & Mrvar, 1998), or Python libraries, whose integration might bring significant contributions to the field of psychometric network analysis.

## Supporting information

Selected Papers for Review

## Data Availability

All data produced in the present work are contained in the manuscript

## Appendix A

### Historical Recap of the Network Approach to Psychopathology

The network approach to psychopathology has been only recently applied to the investigation of eating disorders. However, the theoretical setting of this new perspective already started to be discussed a couple of decades ago.

In the early 2000s, Nancy S. Kim and Woo-Kyoung Ahn already showed that clinical psychologists are cognitively driven to interpret symptom patterns in terms of causal networks (Kim & Ahn, 2002). A few years later, inspired by the philosopher Richard Boyd, the psychiatrist Kenneth Kendler suggested abandoning the traditional disease model in favor of a new quest for complex and multi-level causal mechanisms that produce, underlie, and sustain psychiatric syndromes (Kendler, 2008; Kendler et al., 2011). While answering the question of what kinds of things are psychiatric disorders, he argued that they are most likely to be “*mechanistic property clustersr*” (MPC) kinds, which is to say, rather than having a deterministic essence, they can be described by mutually reinforcing networks of causal mechanisms (Kendler et al., 2011).

In the same period, another critique to a diagnostic system based on the latent factor model was moved by Borsboom (2008). He argued that one important consequence of this model is that one should assume that the property of local independence holds, which is to say, the covariance among symptoms should vanish upon conditionalizing on the presence of the disorder. Nevertheless, as many studies suggest, not only most psychiatric disorders do not satisfy the property of local independence, but also the *Diagnostic and Statistical Manual of Mental Disorders* (DSM) criteria often specify direct functional relations between signs and symptoms (McNally, 2021). Hence, the author put forward the hypothesis of an alternative view of mental disorders as causal systems (Borsboom, 2008).

Later, in order to give a more plausible explanation to comorbidity in the field of psychopathology compared to the latent factor theory, the Psychological Methods program group of the of the Psychology Research Institute of the University of Amsterdam started to develop a network approach to mental disorders and comorbidity in which symptoms are viewed as components in a network and comorbidity is hypothesized to arise from direct relations between symptoms of multiple disorders (Borsboom, 2017; Borsboom & Cramer, 2013; Borsboom et al., 2011; Borsboom et al., 2021; Cramer et al., 2010).

From that point on, more and more studies have been conducted by the same research group and others not only to lick the theory behind the network approach to psychopathology into shape, but also to refine and widen the required methodologies (Brooks et al., 2020; Costantini et al., 2015; Costantini et al., 2019; Epskamp, Borsboom, et al., 2018; Letina et al.) and to explore its applications to some specific mental disorders (Borsboom, 2017; McNally, 2021).

## Appendix B

### Clinical Implications of the Network Approach to Psychopathology

The conceptualization of the network approach to mental disorders does not limit its influence as should not be regarded as a theoretical finding only. Indeed, it has remarkable implications for the diagnosis and treatment systems as well (Borsboom, 2017). With respect to diagnosis, the network approach suggests clinicians to follow a two-step process according to which they should firstly identify the exhibited symptoms and then the network interactions that sustain them, which however is actually not very different from the current DSM diagnostic practice (Borsboom, 2017); the novelty introduced by this approach is that it can help recognize the most important symptoms as well as other significant signs that are not included in the DSM criteria (Fried et al., 2016), such as feeling ineffective and social insecurity in case of EDs.

Finally, as for the disorder treatment, the network perspective suggests clinicians to apply techniques that can change or manipulate the network, in particular that are capable of: changing the state of one or more symptoms, modifying the external field by removing triggering events or manipulating the network structure by altering the connections among symptoms (Borsboom, 2017)

## Footnotes

1 *Recovery Record* is an eating disorder management app used for either patient or clinicians (Tregarthen et al. 2015).

2 The Markov Condition (MC) for a given DAG G with vertex set V and probability distribution P over V states that, conditional on the set of all its direct causes, each node in V is independent of all variables which are not direct causes.

3 A latent confounder is an unmeasured variable that casually influences two or more measured variables

4 Ecological Momentary Assessment (EMA) is a tool designed to collect time-series data from repeated sampling of an individual’s behaviors in their natural environment. Usually, data are collected through an electronic diary or smartphone app that, at given time points, alerts the user to complete some assessment questions and sends the results to the evaluator (Luiselli & Fischer, 2016; Stone & Shiffman, 1994)

5 Multidimensional Assessment of Interoceptive Awareness (MAIA): self-report questionnaire that measures eight facets of interoceptive body awareness (Mehling et al., 2012).

6 Depression, Anxiety, and Stress Scales - Short Version (DASS-21): self-report instrument designed to measure the three related negative emotional states of depression, anxiety and tension/stress, short version composed of 21 items (Lovibond & Lovibond, 1996; Henry & Crawford, 2005)

7 Movie for the Assessment of Social Cognition (MASC): sensitive video-based test for the evaluation of subtle mind-reading difficulties (Dziobek et al., 2006).

8 Empathic Accuracy Task - Revised (EAT-R): test aimed at capturing dynamic aspects of empathy by using videoclips in which perceivers continuously judge emotionally charged stories (Coll et al., 2017; Mackes et al., 2018)

9 Frost Multidimensional Perfectionism Scale (FMPS): self-report measure with four subscales of perfectionism (Frost et al., 1990)

10 Physicia’s Health Questionnaire-Depression Module (PHQ-9): it measures depression symptom severity (Kroenke et al., 2001)

11 Social Interaction Anxiety Scale (SIAS)/Social Phobia Scale (SPS)-Short Forms: they measure generalized social interaction anxiety and specific fears of social scrutiny (Fergus et al., 2012).

12 Structured Clinical Interview for DSM-5 (SCID-5): semi-structured interview used to make DSM-5 diagnoses (First et al., 2015).

13 Positive and Negative Affect Schedule (PANAS): 20-item self-report measure of negative and positive affect (Watson, et al., 1988).

14 Metacognition Self-Assessment Scale (MSAS):18-item self-report questionnaire that evaluates metacognitive functioning, specifically monitoring, differentiation/decentration, integration and mastery (Pedone et al., 2017).

15 Difficulties in Emotion Regulation Scale (DERS): 36-item scale that assesses emotion dysregulation across six subscales, namely, non-acceptance of emotions, difficulties in pursuing goals when having strong emotions, difficulties in controlling impulsive behaviors when experiencing negative emotions, lack of emotional awareness, limited access to emotion regulation strategies, and lack of emotional clarity (Giromini et al., 2012).

16 Youth Self Report (YSR): 112-items self-report questionnaire covering nine syndrome subscales, that is, withdrawn, somatic complaints, anxious/depressed, delinquent behavior, aggressive behavior, social problems, attention problems, obsessive– compulsive problems, and posttraumatic stress problems (Achenbach & Rescorla, 2007)

17 Tridimensional Personality Questionnaire (TPQ): personality test devised to measure three dimensions of the personality: novelty seeking, harm avoidance and reward dependence (Cloninger et al., 1991).

18 Outcome Questionnaire (OQ-45): measurement of general psychopathology (Jong et al., 2008).

19 Mental Health Continuum Short Form (MHC-SF): 14-items scale that measures overall, emotional, psychological, and social well-being (Lamers et al., 2011).

20 IDentity and EAting disorders (IDEA): self-reported questionnaire that assesses abnormalities in lived corporeality and personal identity (Stanghellini et al., 2012)

21 Childhood Trauma Questionnaire (CTQ): 28-items questionnaires to investigates a self-report recall of childhood maltreatment (CM) experience and differentiates five types of CM, namely, emotional neglect, emotional abuse, sexual abuse, physical neglect, and physical abuse (Bernstein et al., 2003).

22 PTSD Checklist for DSM-5 (PCL-5): 20-item self-report measure used to assess the severity of DSM-5 posttraumatic stress disorder symptoms over the past 30 days. It includes four subscales: reexperiencing, avoidance, NACM, and hyperarousal (Weathers et al., 2013).

23 Young Schema Questionnaire (YSQ): it consists of 205 items divided into 16 subscales corresponding to the 16 early maladaptive schema scales (Young et al., 2003).

24 Rosenberg Self-Esteem Scale (RSES): it assesses global self-esteem (Shapurian et al., 1987)

