## Supplementary material for "Network-Based Methods for Psychometric Data of Eating Disorders: A Systematic Review": Selected Papers for Review

**Supplementary Materials**

**Selected Paper List**

14. Elliott, H., Jones, P. J., & Schmidt, U. (2020). Central symptoms predict posttreatment outcomes and clinical impairment in anorexia nervosa: A network analysis. *Clinical Psychological Science*, *8*(1), 139-154. https://doi.org/10.1177/2167702619865958
15. Forbush, K. T., Siew, C. S. Q., & Vitevitch, M. S. (2016). Application of network analysis to identify interactive systems of eating disorder psychopathology. *Psychological Medicine*, *46*(12), 2667-2677. https://doi.org/10.1017/S003329171600012X
16. Forrest, L. N., Jones, P. J., Ortiz, S. N., & Smith, A. R. (2018). Core psychopathology in anorexia nervosa and bulimia nervosa: A network analysis. *International Journal of Eating Disorders*, *51*(7), 668-679. https://doi.org/10.1002/eat.22871
17. Forrest, L. N., Perkins, N. M., Lavender, J. M., & Smith, A. R. (2019). Using network analysis to identify central eating disorder symptoms among men. *The International Journal of Eating Disorders*, *52*(8), 871–884. https://doi.org/10.1002/eat.23123
18. Forrest, L. N., Sarfan, L. D., Ortiz, S. N., Brown, T. A., & Smith, A. R. (2019). Bridging eating disorder symptoms and trait anxiety in patients with eating disorders: A network approach. *International Journal of Eating Disorders*, *52*(6), 701-711. https://doi.org/10.1002/eat.23070
19. Giles, S., Hughes, E. K., Fuller-Tyszkiewicz, M., Treasure, J., Fernandez-Aranda, F., Karwautz, A. F. K., Wagner, G., Anderluh, M., Collier, D. A., & Krug, I. (2022). Bridging of childhood obsessive-compulsive personality disorder traits and adult eating disorder symptoms: A network analysis approach. *European Eating Disorders Review*, *30*(2), 110–123. https://doi.org/10.1002/erv.2885
20. Goldschmidt, A. B., Crosby, R. D., Cao, L., Moessner, M., Forbush, K. T., Accurso, E. C., & Le Grange, D. (2018). Network analysis of pediatric eating disorder symptoms in a treatment-seeking, transdiagnostic sample. *Journal of Abnormal Psychology*, *127*(2), 251–264. https://doi.org/10.1037/abn0000327
21. Hagan, K. E., Matheson, B. E., Datta, N., L'Insalata, A. M., Onipede, Z. A., Gorrell, S., Mondal, S., Bohon, C. M., Le Grange, D., & Lock, J. D. (2021). Understanding outcomes

in family-based treatment for adolescent anorexia nervosa: a network approach. *Psychological Medicine*, 1–12. https://doi.org/10.1017/S0033291721001604

28. Levinson, C. A., Vanzhula, I., & Brosof, L. C. (2018). Longitudinal and personalized networks of eating disorder cognitions and behaviors: Targets for precision intervention a proof of concept study. *International Journal of Eating Disorders*, *51*(11), 1233-1243. https://doi.org/10.1002/eat.22952
29. Levinson, C. A., Vanzhula, I. A., Smith, T. W., & Stice, E. (2020). Group and longitudinal intra-individual networks of eating disorder symptoms in adolescents and young adults at-risk for an eating disorder. *Behaviour Research and Therapy*, *135*, 103731. https://doi.org/10.1016/j.brat.2020.103731
30. Levinson, C. A., Zerwas, S., Calebs, B., Forbush, K., Kordy, H., Watson, H., Hofmeier, S., Levine, M., Crosby, R. D., Peat, C., Runfola, C. D., Zimmer, B., Moesner, M., Marcus, M. D., & Bulik, C. M. (2017). The core symptoms of bulimia nervosa, anxiety, and depression: A network analysis. *Journal of Abnormal Psychology*, *126*(3), 340-354. https://doi.org/10.1037/abn0000254
31. Liebman, R. E., Becker, K. R., Smith, K. E., Cao, L., Keshishian, A. C., Crosby, R. D., Eddy, K. T., & Thomas, J. J. (2021). Network Analysis of Posttraumatic Stress and Eating Disorder Symptoms in a Community Sample of Adults Exposed to Childhood Abuse. *Journal of Traumatic Stress*, *34*(3), 665–674. https://doi.org/10.1002/jts.22644
32. Mares, S. H. W., Burger, J., Lemmens, L. H. J. M., van Elburg, A. A., & Vroling, M. S. (2021). Evaluation of the cognitive behavioural theory of eating disorders: A network analysis investigation. *Eating Behaviors*, *44*, 101590. https://doi.org/10.1016/j.eatbeh.2021.101590
33. Martini, M., Marzola, E., Brustolin, A., & Abbate-Daga, G. (2021). Feeling imperfect and imperfectly feeling: A network analysis on perfectionism, interoceptive sensibility, and eating symptomatology in anorexia nervosa. *European Eating Disorders Review*, *29*(6), 893-909. https://doi.org/10.1002/erv.2863
34. Meier, M., Kossakowski, J. J., Jones, P. J., Kay, B., Riemann, B. C., & McNally, R. J. (2020). Obsessive–compulsive symptoms in eating disorders: A network investigation.

*International Journal of Eating Disorders*, *53*(3), 362-371. https://doi.org/10.1002/eat.23196

### Computer Code

```
graph = “cor”

graph = “concentration”

graph = ”glasso”
```
